# Long-term waning of vaccine-induced immunity to measles in England

**DOI:** 10.1101/2024.04.18.24306028

**Authors:** Alexis Robert, Anne M Suffel, Adam J Kucharski

## Abstract

**Background:** The proportion of double vaccinated cases during measles outbreaks in England has increased since 2010, especially among teenagers and young adults. Possible explanations include: rare infections in vaccinated individuals who did not gain immunity upon vaccination, made more common as the proportion of the population born before vaccination decreases; or waning of vaccine-induced immunity, which would present new challenges for measles control in near elimination settings.

**Methods:** To assess explanations for observed dynamics, we used a mathematical model stratified by age group, region and vaccine status, fitted to case data reported in England from 2010 to 2019. We evaluated whether models with or without waning were best able to capture the temporal dynamics of vaccinated cases in England.

**Findings:** Only models with waning of vaccine-induced immunity captured the number and distribution by age and year of vaccinated cases. The model without waning generated more single-vaccinated cases, and fewer double-vaccinated cases above 15 years-old than observed in the data (median: 73 cases in simulations without waning, 202 in the data, 187 when waning was included). The estimated waning rate was slow (95% credible interval: 0.036% to 0.044% per year in the best fitting model), but sufficient to increase measles burden because vaccinated cases were almost as likely to cause onwards transmission as unvaccinated cases (95% credible interval for risk of onwards transmission from vaccinated cases was only 7% to 21% lower relative to unvaccinated cases).

**Interpretation:** Measles case dynamics in England is consistent with waning of vaccine-induced immunity. Since measles is highly infectious, a slow waning leads to a heightened burden, with an increase in the number of both vaccinated and unvaccinated cases. Our findings show that the vaccine remains protective against measles infections for decades, but breakthrough infections are increasingly likely for individuals aged 15 and older.

**Funding:** National Institute for Health Research; Wellcome Trust.

**Research in context:** *Evidence before this study:* We searched PubMed up to February 29, 2024, with no language restrictions using the following search terms: (measles) AND (“secondary vaccine failure” OR waning) AND (antibody OR “vaccine effectiveness”), and excluded studies that focused on waning of maternal antibodies in infants. We found evidence of waning of antibody concentration in young adults from laboratory data, but this may not translate into a loss of protection against infection. We also found estimates of vaccine effectiveness per age group from statistical analysis that used the total number of cases across various outbreaks rather than transmission dynamics. We did not identify any study estimating waning rate of measles vaccine from recent measles case dynamics.

*Added value of this study:* Our study uses measles case data from England, reported between 2010 and 2020. We show that the transmission dynamics in that time period was consistent with a waning of vaccine-induced immunity, making infection in young adults more common. We estimated that transmission from vaccinated cases was only slightly less common than transmission from unvaccinated cases. The increase in vaccinated cases and transmission from vaccinated cases increased the burden of measles in near-elimination settings.

*Implications of all the available evidence:* Our study shows that measles cases caused by waning of immunity are becoming more common. As the proportion of the overall population vaccinated against measles increases, and vaccine coverage dropped in many countries near elimination between 2020 and 2022, large outbreaks become more likely. Close monitoring of double-vaccinated cases is needed to assess their ability to cause onward transmission.

## Introduction

Measles vaccines are highly protective against infection^1^, and led to a great decrease in the global burden of measles since the start of immunization programs in the 1970s and 1980s. The probability of primary vaccine failure, whereby individuals did not respond immunologically to the vaccine, is below 5%. The existence of secondary vaccine failure, i.e. the loss of immunity over time after vaccination, or waning of vaccine-induced immunity, was highlighted following the implementation of one-dose routine vaccination programs^2^, and was one of the factors leading to the implementation of a second MMR dose in immunization schedule^3,4^.

Following successful routine immunisation programs, countries in Europe, the Americas, and Asia have become eligible for elimination status since 2000. Measles resurgence between 2015 and 2020 led to outbreaks in these settings and highlighted new routes of transmission, affecting teenagers and young adults^5^. This resurgence was mostly reported in under-immunised communities and linked to past variations in vaccine coverage^6^.

Occasional outbreaks were also reported in highly vaccinated groups^7,8^, leading to concerns over waning of measles immunity among adults vaccinated during their childhood^9^. Immunological studies from Canada^10^, Japan^11^, and Czechia^12^ pointed towards waning of antibodies in young adults who had been vaccinated more than 20 years prior, while no decrease was observed in previously infected individuals^13^. Young vaccinated adults had little exposure to measles in near-elimination settings, showing that waning of vaccine-induced immunity may be related to the time since the end of endemic transmission^14^. However, low levels of antibody concentrations may not result in a complete absence of protection against infection.

Analyses from outbreak data have suggested a drop in vaccine effectiveness (VE) among young adults in France (from 99.6% post-vaccination to 96.7% 16 years after vaccination) and in Berlin (from 99% post-vaccination to 90.9% in 31-40 year-olds)^15,16^. Both studies computed the age-stratified VE using the screening method, a statistical calculation of VE similar to a case-control study where the vaccine coverage in the whole population is considered as the control. Franconeri et al^15^ showed that the VE estimates were sensitive to assumptions on the level of infection-induced immunity: VE in older age groups increased as they added infection-induced immunity. The method implemented in both studies relied on total case number per age group, and did not take into account outbreak dynamics.

Given these observations, it is crucial to understand whether measles case dynamics observed in settings with high vaccine coverage result from a waning of vaccine-induced immunity, or whether changes in the immunity landscape are driving the distribution vaccine status among cases, as fewer adults were born in an era of endemic transmission. We implemented a mathematical transmission model stratified by age, region and vaccine status. Such models are better able to capture the non-linear interplay between vaccination and infection-induced immunity and disentangle its impact on the case data compared to statistical analyses.

We applied this model to measles case data by region and age group in England. Measles in England follows a typical near-elimination transmission dynamics, with sporadic localised outbreaks, and high national vaccine coverage: after large outbreaks between 2011 and 2013, England reached measles elimination status following low levels of transmission until 2017. A resurgence of measles was observed from 2017 onwards^17^.

We modelled three possible explanations for observed dynamics: no waning of immunity; waning depending only on age (i.e. VE starts decreasing when individuals turn 5); and waning depending on age and time since measles stopped being endemic (i.e. same as previously, except for individuals vaccinated before 1990, for whom waning starts in 1990). In this last scenario, waning is impacted by low levels of measles incidence in the country^14^. To assess the most plausible scenario, we fitted the three models to measles case data reported in England between 2010 and 2019, and compared the resulting performance.

## Material and Methods

### Measles case data show an increase in the proportion of vaccinated cases

Data on all confirmed measles cases in England between 2010 and 2019 were collected by Public Health England (now UK Health Security Agency). This dataset includes the date of symptom onset, region of residence, age and vaccine status of each case. Only cases reported in England with no missing information were considered, leading to 7,504 cases (Figure 1). If the region of residency was not reported (996 cases), we used the region of the general practitioner who reported the case.

**Figure 1.**
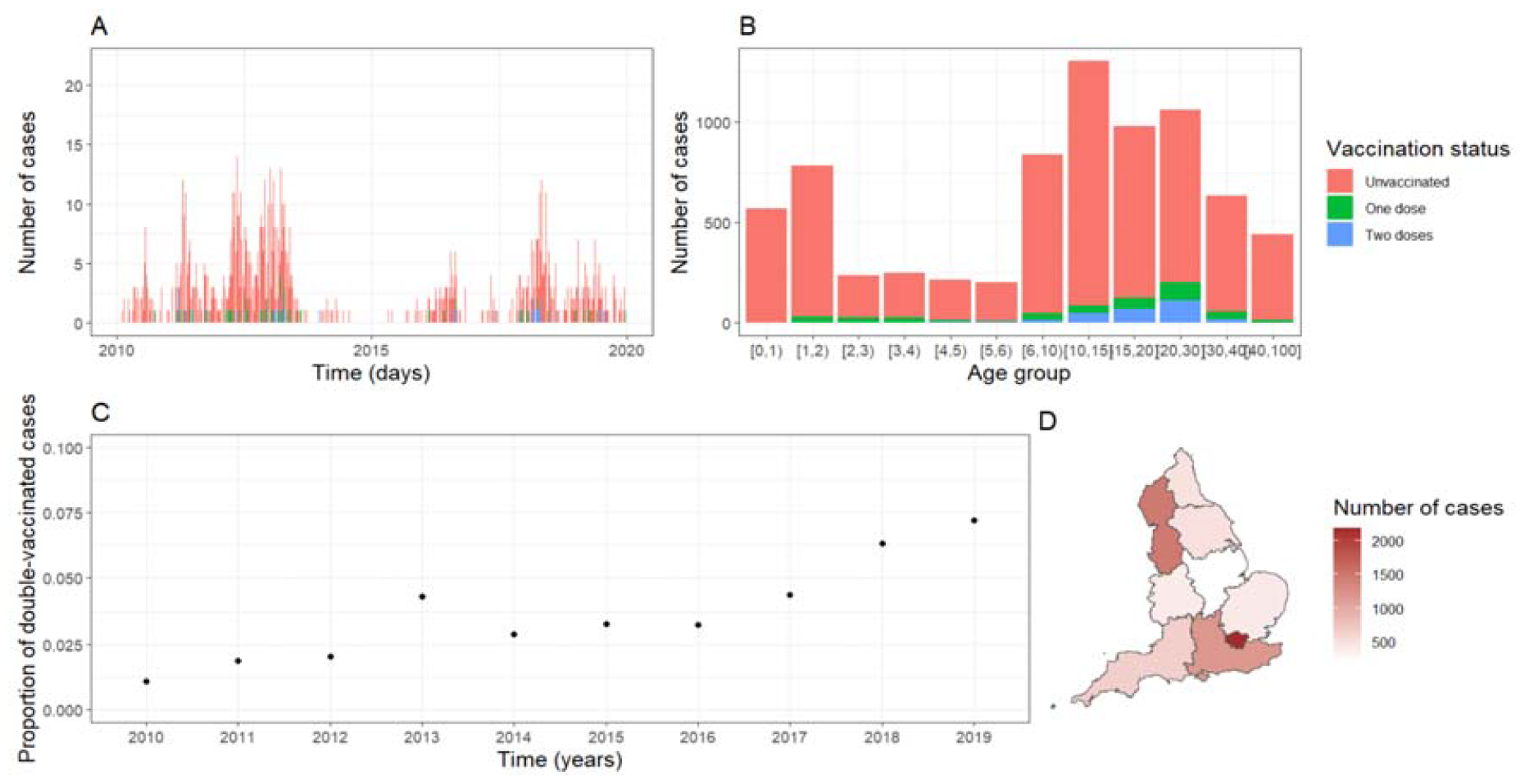
A. Number of daily measles cases reported in England, stratified by vaccine status. B. Number of cases reported between 2010 and 2020 by age group and vaccine status. C. Proportion of double vaccinated cases per year. D. Measles incidence by region of England between 2010 and 2019.

The vaccine status was reported for all cases, labelled “no” for unvaccinated cases, “v1” for single-vaccinated cases, or “v2” for double-vaccinated cases. Between 2014 and 2019, the vaccine status of 143 cases was reported as “yes”. We classified these cases as single vaccinated since no case was otherwise classified as “v1” between 2014 to 2019.

The proportion of double vaccinated cases was three times higher in 2019 than in 2011 (Figure 1, Panels A and C). Most cases were younger than 2 (1,351 cases, 18%), teenagers (2,280 cases, 30.4%), or young adults (1,059 20-30 cases, 14.1%) (Figure 1, Panel B).

### General framework of the compartmental model

We use a compartmental SEIR-type model to fit the number of daily cases per age group, region, and vaccine status in England between 2010 and 2019 (Figure 2). Upon infection, individuals move from “Susceptible” to “Exposed”, from “Exposed” to “Infectious” at the end of the incubation period, and move to the “Recovered” compartment when the infectious period ends.

**Figure 2.**
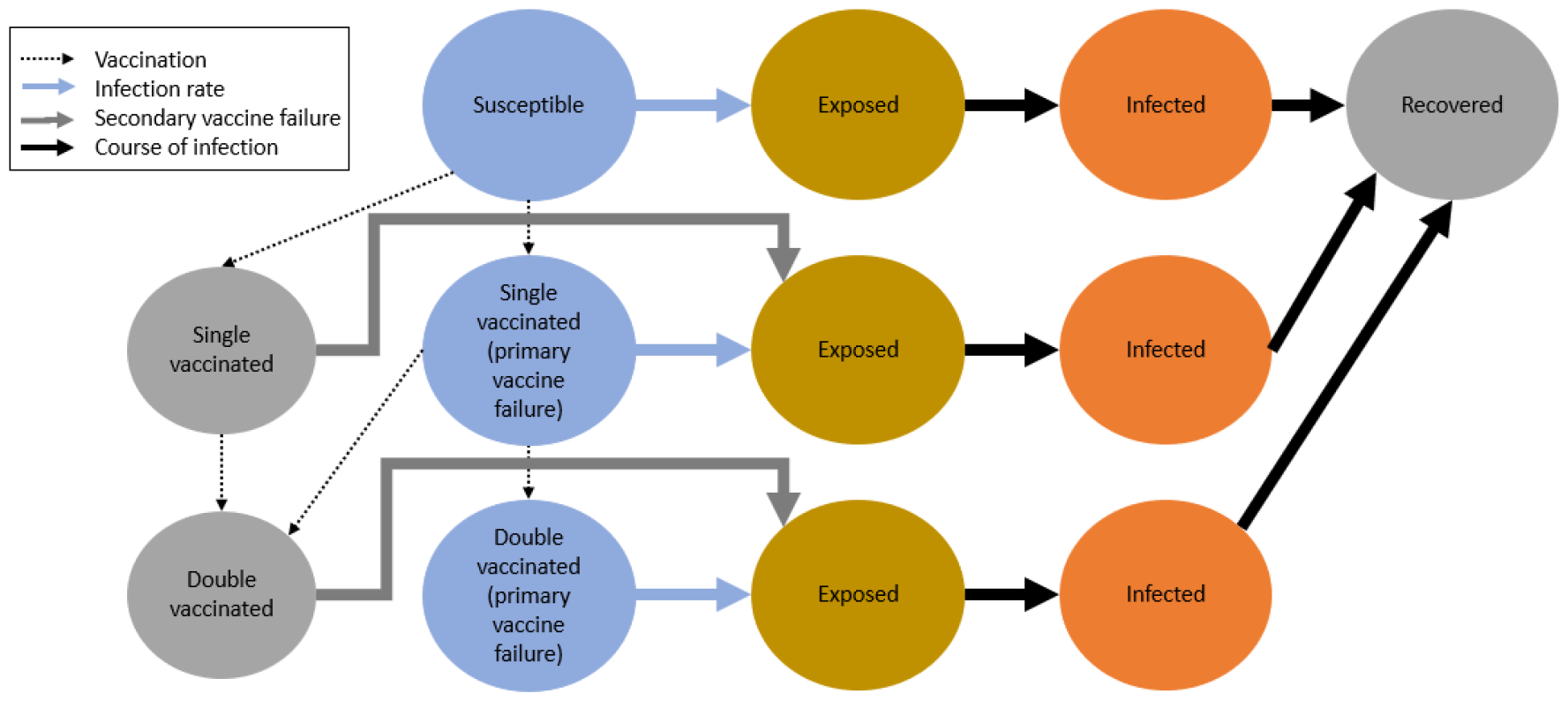
Model schematics for a given age group / region. In age group 0-1, individuals are placed in the Maternal immunity compartment (not pictured) when they are born, and move into the Susceptible compartment, at a rate defined by the duration of maternal immunity. In the age group 40+, individuals exit the Recovered compartment, at a rate corresponding to the number of deaths per day.

We implemented the model using the R package odin.dust. For each strata (age group, region, and vaccine status), the likelihood is computed by comparing the data to the number of cases moving from the Exposed to the Infected compartments each day. The log-likelihood in each strata are then added to compute the overall log-likelihood (Supplementary Section S1).

The model requires parameters quantifying how individuals move between compartments (incubation and infectious periods, which are fixed), how quickly measles spreads (infections rate, seasonality, and contact matrix between age groups and regions), how often measles is imported (number and seasonality of importations), and how protective and effective the vaccine is (rate of primary vaccine failure, protection against onward transmission, and waning rate). The model requires the immune landscape and vaccine distribution of each age group and region of the population at the start time (1^st^ January 2010). All parameters are summarised in Supplementary Section S1 and Supplementary Table S1.

### Vaccination data

The model requires the proportion of single and double vaccinated individuals for each age group, region and year. Two sources of vaccination data were used: Cover of Vaccination Evaluated Rapidly (COVER), a dataset published by NHS Digital summarising UK vaccination coverage at the age 2 and 5 per region for the MMR vaccine^18^ ; and Clinical Practice Research Datalink (CRPD) Aurum, a primary care dataset from GP practices containing patient-level information on immunisations^19^.

Supplementary Section S2 shows a comparison between COVER data and CPRD data. CPRD coverage tends to be higher than raw COVER data, and close to COVER data adjusted for 50% under-ascertainment (i.e. half the unvaccinated individuals were actually vaccinated).

In the reference scenario, data for years not covered by CPRD data (children born before 2006 or after 2015) were supplemented with estimated values by region based on COVER data adjusted for under-ascertainment. These corrected values were consistent with estimates from previous studies^20^.

Region-stratified coverage data was not available for children born before 2004, so we used the first and second dose coverage from UKHSA’s Risk assessment for measles resurgence in the UK, which gives the national coverage in the country and in London^21^.

In England, the first MMR dose is given from 1 year of age, and the second from 3 years and 4 months. First-dose coverage at one was set to 75% of coverage at two in the model, so children would not be fully susceptible until they reach 2 years of age. Similarly, we set the second-dose coverage at three to 50% of the coverage at four. We assumed that no individual in the 30-40 and over 40-year-old categories in 2010 had been vaccinated.

### Models and sensitivity analysis

We compared the outputs from three models: without waning of immunity, with waning of immunity increasing each year from 5 years old, with waning of immunity increasing each year from 5 years old and from 1990. In this last scenario, individuals vaccinated before 1990 have full protection until 1990, waning of immunity is then linked to low levels of transmission observed since 1990 in England^22^.

We used a deterministic framework to fit the model to the data, we compared the posterior distributions to find the best performing model. The parameter sets obtained from model fits were used to generate stochastic simulations, showing the transmission dynamics generated by the model between 2010 and 2019. Fitting a stochastic version of the model was not computationally feasible given the number of compartments and strata in the model. Additional figures describing the reference scenario fits and simulations are shown in Supplementary Section S3.

We explored various sensitivity analyses to assess whether the impact of waning of immunity was robust to changes in assumptions:

- Addition of constant risk of secondary vaccine failure estimated by the model: the protection against infection is not perfect, but is not waning over time (Supplementary Section S4).
- Using the COVER data without adjustment for under-ascertainment, and use the CPRD data to compute the proportion of new vaccinated at 3 and 4 (Supplementary Section S5).
- Set the values of the risk of cross-regional transmissions to test different transmission patterns between regions (Supplementary Section S6).

## Results

### Only models with waning of vaccine-induced immunity capture the transmission dynamics in vaccinated cases

Models with waning had a better posterior distribution than the model with primary vaccine failure only (Supplementary Figure S3), as they were better able to capture the distribution of vaccinated cases. When waning of immunity was not included in the model, the simulations over-estimated the number of single vaccinated cases (Median 561 [95% simulation interval (SI): 354-939] cases, 362 cases in the data), and under-estimated the number of double-vaccinated cases (Median 168 [95%SI: 102-287] cases, 277 cases in the data) (Figure 3). Similarly, the model without waning overestimated the number of single and double vaccinated cases among children (5-15 years old), and greatly underestimated the number of double vaccinated among teenagers and adults (Median 73 [95%SI: 44-121] cases, 202 cases in the data) (Figure 3, Panels B-G). When incorporating a constant risk of secondary vaccine failure, the model without waning captured the overall number of both single and double vaccinated cases (Supplementary Section S4), but did not capture their age distribution (Supplementary Figure S9). Only simulations generated with models including waning of vaccine-induced immunity captured the age distribution of the vaccinated cases.

**Figure 3.**
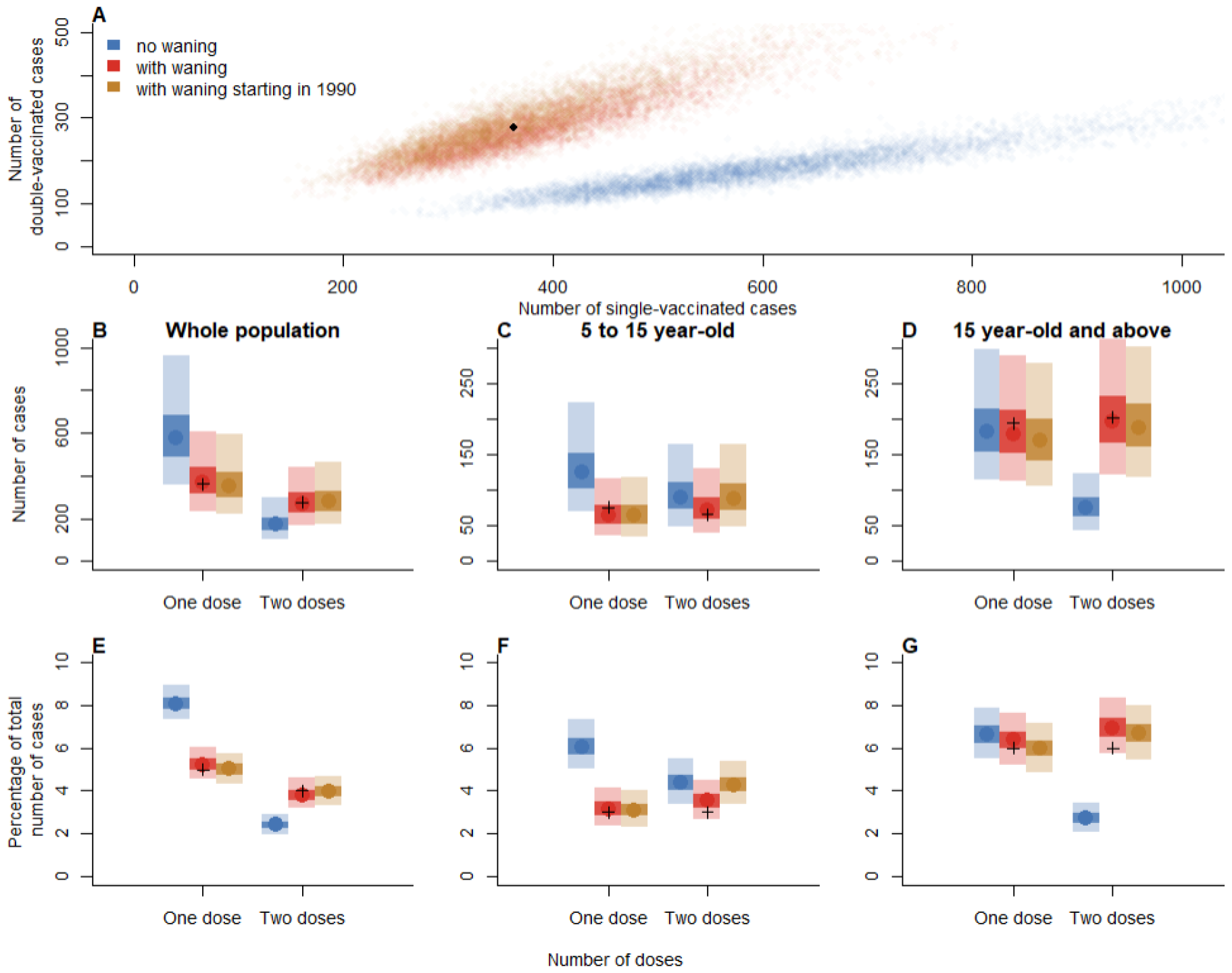
A. Comparison of the number of single and double vaccinated cases in models with and without waning (orange and red dots cover the same area, so may be masking each other), the distribution in the data is shown by a black dot. B. Overall number (and E. Proportion) of single and double vaccinated cases in each model (data points are represented by black crosses). C. Number (and F. Proportion) of vaccinated cases between 5 and 15 years old. D. Number (And G. Proportion) of vaccinated cases above 15 years old. All plots show the number of cases across all regions and years.

The distribution of vaccinated cases through time was also better captured by models incorporating waning of immunity (Figure 4). The proportion of single vaccinated cases per year was consistently over-estimated in simulations that did not include waning of immunity. The simulations generated with both models with waning captured the number of single-vaccinated cases every year but in 2015, where case numbers were low (92 cases in 2015, 1 single-vaccinated). When waning of immunity was included, the proportion of double-vaccinated cases increased through time. However, this increase was slower than the one observed in the data. When waning started in 1990, the increase was slightly faster, and the posterior distribution slightly better than in the baseline model with waning (Figure 4, Panel B, and Supplementary Figure S3).

**Figure 4.**
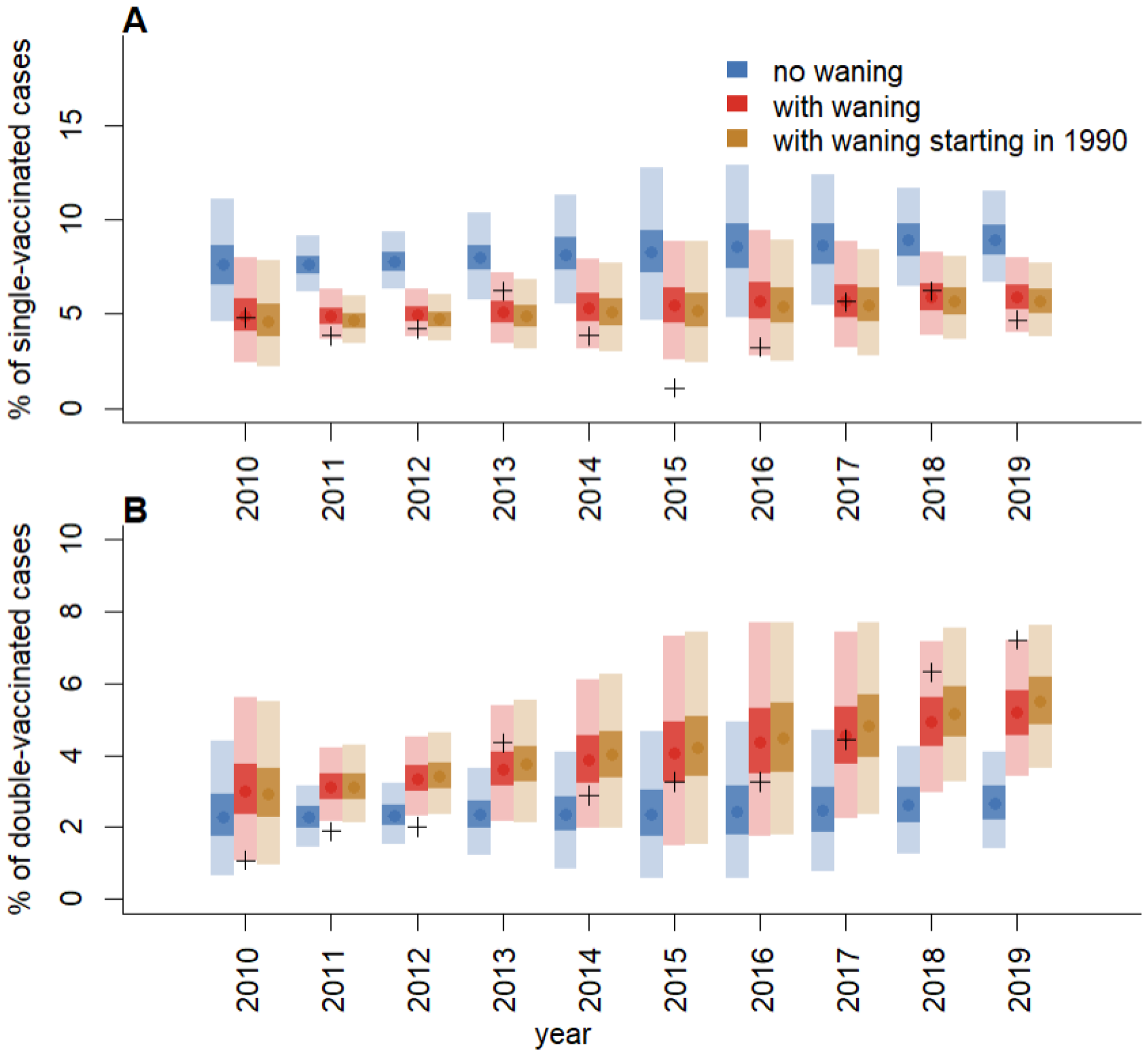
A. Proportion of single (and B. double) vaccinated cases each year across all regions and age groups.

### Waning leads to higher levels of transmission

In all models, R0 was estimated between 16 and 18 (Figure 5, Panel A, Supplementary Figure S5, Panel C)., on the higher side of the typical R0 estimates for measles^23^. Models incorporating waning of immunity estimated a decrease in VE through time, although VE remained high after several decades (Figure 5 Panel D). The waning rate was 0.039% per year (95% credible interval (CI): 0.034-0.044%) in the model with waning starting in 1990. The rate of primary vaccine failure was higher in the model without waning (median estimate 5% without waning, 2% otherwise) (Figure 5, Panel C and Supplementary Figure S5, panel F). In the reference scenario, the risk of onward transmission from vaccinated cases compared to unvaccinated cases was 82% (95CI: 72-91%) (Figure 5, Panel B), so vaccinated cases were almost as likely to cause secondary transmission as unvaccinated cases. This risk was lower when using the COVER data (between 10 and 45%, Supplementary Figure S16).

**Figure 5.**
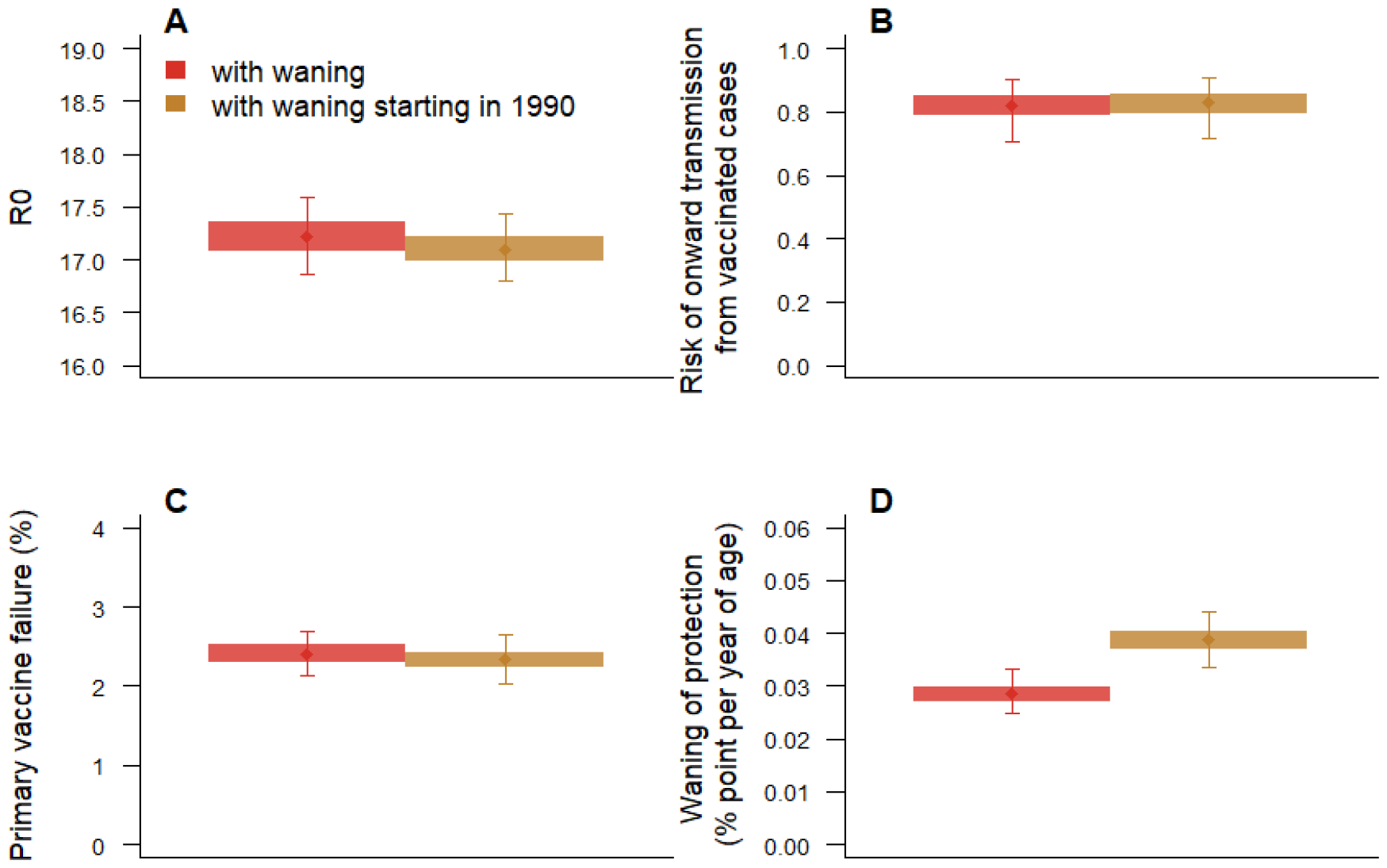
Key parameter estimates in the best fitting models with waning of vaccine-induced immunity (using CPRD vaccine data). A. Basic reproduction number R_0_. B. Risk of onward infection from vaccinated cases compared to unvaccinated cases. C. Proportion of primary vaccine failure. D. Rate of waning of vaccine-induced immunity (in percentage point per year of age).

We estimated the impact of waning on case numbers by setting the waning rate to 0 in models that incorporate waning, and comparing case numbers with the reference simulations (Supplementary Figure S8). In models using CPRD data, removing waning substantially decreased the number of cases, especially in 2018 and 2019 (210 to 728 cases in the simulations in 2018, 963 in the data; 252 to 824 cases in the simulations in 2019, 790 in the data). However, when using COVER data, removing waning had less impact (Supplementary Figure S17).

The overall distribution of cases by age group, year or region was the same in all models (Supplementary Figure S6). The number of cases by age groups was in agreement with the data, although the model over-estimated the number of infants reported. The model underestimated the number of cases reported in North West and North East, and overestimated the burden in East and West Midlands. This is expected as R0 was not stratified by region, and spatial heterogeneity in transmission risk only depended on region-stratified vaccine coverage, available for cases vaccinated from 2004 onwards.

## Discussion

We found that only transmission models that included waning of vaccine-induced immunity were able to capture the age and transmission dynamics of vaccinated cases. In the best-performing model, the estimated waning rate was 0.039% per year (95%CI: 0.034-0.044%). Although slow, this waning was associated with an increased burden over time: removing the waning process from the model led to a substantial decrease in cases (58% median reduction compared to 2018 data, 38% decrease in 2019). Although the overall vaccine effectiveness remained high over the decades despite this waning, our estimation suggests the increasing number of breakthrough infections had a measurable effect on observed dynamics.

The additional disease burden brought by waning is directly related to the risk of transmission from vaccinated cases, since individuals infected by vaccinated cases would not have been infected without waning. In the reference scenario, the model estimated that vaccinated and unvaccinated cases had similar rates of onward transmission (Figure 5 Panel B), but this rate was lower in vaccinated cases when using the COVER data (Supplementary Figure S16). Epidemiological reports have showed rare onwards transmission events from vaccinated cases^8,24^, potentially because unvaccinated cases cluster in vulnerable groups so opportunities of transmission are rarer^25,26^. Contact tracing investigations or transmission tree reconstruction methods^27^, with better spatial resolution, should be used to quantify how often vaccinated cases are associated with onward transmission.

Early signs of waning-linked transmissions have been observed through outbreaks in near-elimination settings (Europe^7^, Japan^8^, and United States^28^). Given we show that transmission dynamics in England are consistent with gradual waning of immunity, future analyses should assess whether other near-elimination countries show similar waning rates, and identify population-level factors that may influence the waning rate. Such estimate will be crucial to evaluate our ability to eliminate measles in high-coverage settings. The proportion of double-vaccinated cases per year is increasing faster in the data than in all models, which may indicate waning dynamics more complex than what we tested. The models only integrate linear waning (i.e an absolute reduction in VE each year of age), but age-specific variations in the waning rate may better explain the data^13^.

Epidemiological reports have highlighted that symptoms in vaccinated cases are milder, thereby increasing the risk of underreporting^8,24^. National measles guidelines in the United Kingdom state that the number of vaccinated cases is expected to increase with higher availability of testing and better reporting rate^29^. As higher availability of testing could also increase the number of unvaccinated cases, the proportion of vaccinated cases would then remain unchanged. Improvements in testing patterns for certain settings or subpopulations (e.g. healthcare settings) would lead to increase in case numbers specific to vaccinated cases, especially as unvaccinated cases are more likely to be part of marginalised communities with less access to healthcare and vaccination. This may partly explain the increase in proportion of vaccinated cases. However, only the proportion of double-vaccinated cases increased between 2010 and 2020, and the proportion of single-vaccinated cases was constant (Figure 4), indicating changes in dynamics specific to double-vaccinated adults and teenagers. We did not have access to data on testing per year, age, or region that could be integrated in the model, which would be needed for future analyses looking into the impact of reporting.

In our model, the waning of vaccine-induced immunity is only based on the age of the individuals, assuming that waning starts at age 5. The small proportion of individuals who were vaccinated later in life (e.g. during catch-up campaigns), have the same VE as individuals vaccinated before age 5. Allowing for disparities would have required adding multiple compartments to the model.

The estimates of vaccine coverage rely on data up to age 5, which may not include movements of teenage and adult population, and late vaccination. To account for these limitations and test their impact on the conclusions, we allowed the model to estimate the impact of catchup campaigns pre-2010, and used several vaccine datasets (Supplementary Section S5). In all scenarios, the models without waning were not able to capture the number and age distribution of vaccinated cases.

Within a given age group, region, and vaccine status, the model was homogeneous, ignoring the variations in transmission risk within regions. Measles outbreaks in near-elimination settings are triggered by importations in pockets of susceptibility where vaccine coverage is low, which would not be identified as the spatial granularity in the model is too coarse. Although this assumption makes compartmental models inappropriate to estimate the future risk of outbreaks, we do not anticipate that it would affect the vaccine distribution of the cases or impact the estimates of the waning rate.

To ensure that the parameters in the baseline model can be statistically identified, the infection rate does not depend on the region, or age group. Region and age-specific outbreak risk only depends on the spatial kernel, contact matrix, and vaccine coverage, all of which are only captured by estimations. This assumption leads to discrepancies between the spatial distribution of cases in the data and simulations (Supplementary Figure S6). As a sensitivity analysis, we implemented a version of the model where the parameters of the gravity model are set rather than estimated, and found that models with waning of immunity still better capture the vaccine distribution of the cases (Supplementary Section S6).

Our results show that waning of vaccine-induced immunity best explains the observed dynamics and age distribution of vaccinated measles cases in England. As measles vaccine coverage has decreased in many near-elimination countries since 2020^30^, the risk of outbreaks is high. England has already reported a sharp increase in case numbers in 2023-2024. Accounting for the impact of waning – as well as declining coverage – on future measles dynamics will be paramount to anticipating the burden of measles in countries where incidence has been low for decades.

## Data Availability

The individual-level case data was collected by UKHSA and cannot be shared publicly. The code used to generate the fits, simulations and figures presented in the paper is shared in a Github repository (https://github.com/alxsrobert/measles_england_sir). This repository contains the model fits generated using the case data, the stochastic simulations, and all population and coverage data used in the analysis. In order to make this study as reproducible as possible, we generated a simulated linelist and included it in the Github repository, so readers can generate model fits on the simulated datasets.

## Acknowledgments

We would like to thank Vanessa Saliba from UKHSA for their feedback on the manuscript, and UKHSA for collecting the case data analysed in this study.

## Authors’ contribution

AR, AMF and AJK developed the analysis plan. AR implemented the analysis, wrote the code and ran the model. AMF computed and collated the coverage data. AR and AMF interpreted the results, with contributions from AJK. AR wrote the first draft and the additional file. AR, AMF and AJK contributed to the manuscript, all authors read and approved the final version of the manuscript.

