## Supplementary Material for "Long-term waning of vaccine-induced immunity to measles in England"

### Section S1: Model description and equations

This section describes the stratifications and parameters used in the model, all parameters are described in Supplementary Table S1. The model was implemented using the R package *odin.dust* (1).

#### Infection rate

The infection rate is the same in each region (i.e. $\beta_{london}=\beta_{east of england}=.. =\beta)$, age ($\beta_{0-1}=\beta_{1-2}=.. =\beta)$, and year ($\beta_{2010}=\beta_{2011}=\ldots=\beta$). The rate of onward transmission in a given region is affected by the contact pattern between age groups, the connectivity to other regions, and the proportion of vaccinated and immune individuals in the region. Two parameters $X$ and $Y$ are estimated by the model to quantify the seasonality in infection rate (i.e. how the infection rate changes each day of the year), such that for each time $t$:

$$\beta_{t}=\beta*(X* cos \left( \frac{2*\pi*t}{365.25}+Y \right))$$

Therefore, the model estimates three parameters to quantify the infection rate through time: $\beta$, $X$, and $Y$.

#### Vaccine stratification

The model uses an all-or-nothing vaccination structure to represent primary vaccine failure: given $v_{fail}$ the proportion of vaccine failures, individuals move to one of the Vaccinated compartments upon first vaccination: they have a $v_{fail}$ risk of not being protected against infection (i.e. primary vaccine failure), and $1-v_{fail}$ chances of gaining full protection against infection. When and if individuals who did not get protected get their second dose of vaccine, they have another $v_{fail}$ risk of not gaining protection, and a $1-v_{fail}$ chances of gaining full protection. Individuals are protected if at least one dose did not fail. The risk of primary vaccine failure is the same for each dose.

When waning is added to the model, the “vaccinated and protected” compartments become leaky and are not fully protective anymore. The parameter $v_{leak}$ quantifies the amount of protection lost by the vaccines in absence of primary vaccine failure and is estimated by the model ($p_{protect}=1$ at 5 years of age, and decreases every year after vaccination: $p_{protect}\left( a \right)=1-\left( a-5 \right)*v_{leak}$). We consider that protection wanes at the same rate for single and double vaccinated individuals.

The model also includes $v_{onwards}$, the reduction in onward transmission for vaccinated cases compared to unvaccinated cases. This reduction corresponds to both potential protection from the vaccine, and clustering of vaccinated cases with other vaccinated individuals (i.e. vaccinated cases may be less likely to be in contact with unvaccinated individuals than unvaccinated cases).

#### Age stratification

The model includes 12 age groups: <1, 1-2, 2-3, 3-4, 4-5, 5-6, 6-10, 10-15, 15-20, 20-30, 30-40 and 40+ years old.

##### Ageing and vaccination

We use the vaccine data to compute the number of children moving from S to V1, and from V1 to V2 in each age group at each year:

For a given age group $i$, the number of individuals gaining vaccination as they age into this age group is computed from a binomial distribution $B({nS}_{ageing}, p_{vacc}1)$, with $p_{vacc1}$ the proportion of new single-vaccinated, and ${nS}_{ageing}$ the number of unvaccinated ageing at this time step. With $cov1(a,t)$ the proportion of single-vaccinated at age $a$ and time $t$, and $cov2(a,t)$ the proportion of double-vaccinated at age $a$ and time $t$ The proportion of new single-vaccinated is calculated as:

$$p_{vacc1}\left[ i \right]=1- \left( proportion of unvaccinated who do not get vaccinated as they age into age i \right)=1-\frac{proportion of unvaccinated at age i\mathrm{in}the current year}{proportion of unvaccinated at age (i-1) in the previous year}=1-\frac{1-cov_{1}(i, t)-cov_{2}(i, t)}{1-cov_{1}(i-1, t-365)-cov_{2}(i-1, t-365)}$$

Similarly, the number of new double-vaccinated individuals is drawn from a binomial distribution $B(nV1_{ageing}, p_{vacc2})$ , with $p_{vacc2}$ the proportion of new double-vaccinated, and ${nV1}_{ageing}$ the number of single-vaccinated ageing at this time step. The proportion of single vaccinated who become double-vaccinated is calculated as:

$$p_{vacc2}\left[ i \right]=\left( proportion of single vaccinated who get vaccinated \right)=\frac{proportion of new double vaccinated}{proportion of single vaccinated at previous age and year}=\frac{cov_{2}(i, t)-cov_{2}(i-1, t-365)}{cov_{1}(i-1, t-365)}$$

##### Contact between age groups

The contact pattern between age groups is quantified by a contact matrix, computed from the POLYMOD study. We use the POLYMOD matrix from England to compute the contact rate between age groups per capita(2).

##### Immunity

The model requires the distribution of vaccine and recovery status for each age group and region at the start date of the fit (1^st^ January 2010). We use parameters and vaccine data to estimate the vaccine distribution and the amount of infection-induced immunity for each age group, depending on the data available:

- Individuals born in 2004 and after: We used vaccine data for each region to get the proportion of single and double vaccinated individuals for each age group.
- Individuals born prior to 2004 in London: We used vaccine data in London to get the proportion of single and double vaccinated individuals for each age group.
- Individuals born prior to 2004 outside of London: We used vaccine data at a national level to get the mean coverage of the first and second dose in each age group. Region-stratified data was not available for these years, except for London. Therefore, all regions except London have the same value of coverage.
- Individuals born before 1980 are set as unvaccinated. The proportion of these unvaccinated individuals who had previously been infected is estimated by the model.
- Existing natural immunity: Five parameters are estimated by the model to quantify the proportion of individuals in the R compartments at $t=0$ per age group: $recov_{10-15}$, $recov_{15-20}$, $recov_{20-30}$, $recov_{30-40}$, and $recov_{40+}$. These proportions do not vary by regions. Immunity in children below 10 years old is set to 0.
- Catch-up campaigns: Several vaccine catch up campaigns have been carried out in England between 1995 and 2010:
  - targeting children vaccinated once born in the late 1980s and early 1990s (MMR2 catchup in 1996). This is controlled by the parameter $catchup$, quantifying the proportion of single-vaccinated individuals aged 20 to 30 in 2010 who got their second dose during the catchup campaign. This parameter is the same across all regions.
  - targeting all children under 18s (MMR national catch-up programme in 2008). This is controlled by the parameter $catchup2$, quantifying the proportion of previously single-vaccinated and unvaccinated individuals aged 6 to 10 in 2010 who got vaccinated twice during the catchup campaigns. We consider the impact of this campaign on individuals born between 2000 and 2005 since they were prioritised (“younger children with no MMR doses[..] were prioritised”(3)). This parameter is the same across all regions.

Finally, we compute the duration of maternal immunity (parameter $\delta$) to estimate the proportion of children below one-year-old who are susceptible to infection. New births are placed in the M compartment, where they are fully protected from infection, and can only move to the S compartment (the rate of movement is computed from the duration of the maternal protection).

##### Demography

The number of inhabitants per year and age group in 2010 was computed using the 2006 census. The number of births per year is computed using the population censuses in 2006, 2013, and 2019. The number of inhabitants in each region is considered constant through time: the number of deaths is equal to the number of births, and all deaths come out from the oldest age group (> 40 year old individuals). This simplification ignores various important demographic factors that could affect measles transmission in a country (e.g. migration between regions, migration outside the country, deaths in younger age groups).

#### Region stratification

##### Transmission between region

There are nine government office regions in England: South West, South East, London, East of England, East Midlands, West Midlands, Yorkshire and the Humber, North East, and North West. Although migration of inhabitants is not represented in the model, the model includes a spatial kernel, which quantifies the rate of transmission from one region to another. This kernel is a gravity model, depending on population in both regions, and distance. Distance is accounted for by the degree of connectivity between regions (neighbours have a degree of one, neighbours of neighbours have a degree of two etc.). The spatial kernel is modelled using a gravity model. The rate of transmission between two given regions $k$ and $l$is computed as:

$$d\left( k,l \right)=N_{l}^{b}*\theta*D\left( k,l \right)*\frac{N_{k}^{c}}{N_{k}}$$

With $N_{k}$ and $N_{l}$ the number of inhabitants in regions $k$ and $l$, $D(k,l)$ the distance between $k$ and $l$, and $b$, $c$ and $\theta$ parameters estimated by the model.

Putting the different elements of the model together, the probability of an unvaccinated susceptible individual in a given age group and region being exposed at time $t n(a,i,t)$ is computed as:

$$p\left( a,i,t \right)=1-exp(-\frac{\beta\left( t \right)}{N}*\sum_{j=1}^{j=12} \sum_{l=1}^{l=9} m\left( j,a \right)*d\left( l,i \right)*\left( Is\left( j,l,t \right)+v_{onwards}*\left( Iv1\left( j,l,t \right)+Iv2\left( j,l,t \right) \right) \right))$$

With N the total number of inhabitants, $m$ the contact rate between age groups per capita, and $v_{onwards}$ the protection against onward transmission given by the vaccine.

The probability of individuals who were vaccinated but did not gain protection becoming exposed is the same as unvaccinated individuals ($p\left( a,i,t \right))$.

The probability of individuals “vaccinated and protected“ becoming exposed is 0 in models that do not include waning. In models with waning, the probability of becoming exposed depends on the level of waning at the age of the individual ($p_{protect}\left( a \right)=1-\left( a-5 \right)*v_{leak}$), such that $p_{vaccinated}\left( a,i,t \right)=p\left( a,i,t \right)*p_{protect}(a)$.

#### Importation

The average number of importations per year and region was computed from the number of cases classified as “imported” or “import related” in the individual case data for each region and year. This local number of importations for each year is then divided by 365 and weighted by the number of inhabitants per age group, to get the daily importation rate by age group and region. We consider that importations are less likely to be reported than other cases, so the importation rate by region is divided by $p_{import}$, the probability of reporting of imported cases. The model also estimates two parameters $X_{import}$ and $Y_{import}$, to estimate the seasonality of importations in a given year.

$$n_{import}\left( a,i, t \right)=\frac{n_{import}(i)}{365*p_{import}}*\frac{N_{ai}}{N_{i}}(X_{import}*\cos\left( 2*\pi*\frac{t}{365.25}+Y_{import} \right))$$

With $N_{ai}$ the number of inhabitants of age $a$ in $i$, and $N_{i}$ the number of inhabitants in region $i$.

| Parameter | Fixed / estimated | Value / Prior | Reference |
| --- | --- | --- | --- |
| Infection rate $\beta$ | Estimated | U(0, 50) |  |
| Duration of maternal immunity (days) $\delta$ | Estimated | N(90, 5) | (4) |
| Risk of onwards transmission from vaccinated cases, compared to unvaccinated cases $v_{onwards}$ | Estimated | B(10,10) | (5) |
| Seasonality of infection rate: $X$ | Estimated | U(0, 7) |  |
| Seasonality of infection rate: $Y$ | Estimated | U(0, 7) |  |
| Mean duration of latent period (days) $\gamma$ | Fixed | 11 | (6,7) |
| Mean duration of infectious period (days) $\alpha$ | Fixed | 8 | (7) |
| Waning of immunity per year $v_{leak}$ | Estimated or Fixed | U(0,1) or 0 |  |
| Proportion of primary vaccine failure $v_{fail}$ | Estimated | U(0, 1) |  |
| Risk of secondary vaccine failure $v_{sec}$ | Fixed* | 0 |  |
| Proportion of 20-30 yo individuals vaccinated during the 1996 catch-up campaigns $catchup$ | Estimated | U(0, 1) |  |
| Proportion of 6-10 yo individuals vaccinated during the 2008 catch-up campaign $catchup2$ | Estimated | U(0, 1) |  |
| Proportion of previously infected 10-15 yo $recov_{10-15}$ | Estimated | U(0, 1) |  |
| Proportion of previously infected 15-20 yo $recov_{15-20}$ | Estimated | U(0, 1) |  |
| Proportion of previously infected 20-30 yo $recov_{20-30}$ | Estimated | U(0, 1) |  |
| Proportion of previously infected 30-40 yo $recov_{30-40}$ | Estimated | U(0, 1) |  |
| Proportion of previously infected 40+ yo $recov_{40+}$ | Estimated | U(0, 1) |  |
| Spatial parameter $b$ | Estimated* | U(0, 5) |  |
| Spatial parameter $c$ | Estimated* | U(0, 5) |  |
| Spatial parameter $\theta$ | Estimated* | U(0, 1) |  |
| Proportion of importations reported $p_{import}$ | Estimated | U(0, 1) |  |
| Seasonality of importations $X_{import}$ | Estimated | U(0, 7) |  |
| Seasonality of importations $Y_{import}$ | Estimated | U(0, 7) |  |

**Supplementary Table S1: Summary of the parameters. *Parameters fixed or estimated in sensitivity analyses.**

#### Likelihood and posterior

To fit the models, we used Monte Carlo Markov Chains, with 20,000 iterations and a burnin period of 1,000 iterations.

The likelihood was computed by comparing the number of new cases at each date, age group, region, and vaccine status in the data with the number of individuals entering the corresponding Infected compartment in the model at this date.

We use a Poisson distribution to fit the number of cases generated by the model $X_{aitv}$to the data $Y_{aitv}$ (at age $a$, in region $i$, at time $t$, and vaccine status $v$)

$$Y_{aitv}\sim Poisson(X_{aitv})$$

We then compute the log likelihood by summing the log likelihood of each data point (across time, age groups, regions and vaccine statuses).

$$L_{tot}=\sum_{a} \sum_{i} \sum_{t} \sum_{v} log(P_{poisson}(Y_{aitv}|X_{aitv}))$$

The log posterior is then computed by summing the log-likelihood and the log-prior.

#### Stochastic simulations

In the stochastic version of the model, the number of transitions between compartments is drawn using a binomial distribution and the rate of transition, whereas only the rate of transition is used in the deterministic fits. Stochastic simulations were implemented to explore the range of scenarios generated by the parameter sets and the specification of the model. We generated 5,000 stochastic simulations per scenario.

### Section S2: Vaccine data

#### Description and comparison of COVER and CPRD datasets

Cover of Vaccination Evaluated Rapidly (COVER), a dataset published by NHS Digital summarising UK vaccination coverage at the age 2 and 5 for the MMR vaccine, at a regional level (8). Coverage data was available for children born between 2000 and 2019. Local 2-year coverage was available for children born from 2004. Region-stratified coverage data was not available for children born before 2004, so we used the first and second dose coverage from UKHSA’s Risk assessment for measles resurgence in the UK, which gives the national coverage in the country and in London. However, COVER data does not contain information on coverage at ages 1, 3, and 4, and previous reports have indicated that they may be subject to under-ascertainment (9,10).

We also use the Clinical Practice Research Datalink (CRPD) Aurum, a primary care dataset from GP practices containing patient-level information on symptoms and diagnoses, clinical tests and results, immunisations, prescriptions and referrals to other services (11). In 2022, CPRD Aurum contained data from around 25 million patients and was broadly representative of England by geographical spread, age, sex and ethnicity (12).  Using a validated algorithm to identify vaccination records (13), vaccination coverage at the ages of 1, 2, 3, 4, and 5 years was estimated from the electronic health records of 573,015 children who have been followed up until the age of five. The results were previously published (14). As the CPRD data was only available for children born between 2006 and 2015, this covered all age bands between 0 and 5 in the years 2010 to 2016. Therefore, compared to COVER data, CPRD data contains information on how coverage changes at each age group, but covers a more narrow time period.

Vaccination coverage estimates for England in CPRD are consistently between 2 and 4% higher than the COVER estimates (Table S2). The same is true for coverage at regional level, for both first and second MMR dose, the coverage in CPRD usually remains above the coverage in COVER (figures S1 and S2). The only exception is London which has a lower coverage for the second MMR dose in CPRD than in COVER from 2015 onwards (see figure S2). Other temporal trends remain similar across regions in both data sets (figures S1 and S2).

Differences in the coverage estimates can result from different population denominators in both data sets, and GP practices using different IT systems feeding into the datasets. As quality of prescribing data for vaccines also varies by IT system used in the practice and as EMIS (the IT system CPRD is built on) does not provide an extra in-built feature for recording vaccination, the prescription data might be more inaccurate than in data based on other IT systems. Hence, the clinical codes used to extract vaccination data from CPRD were less conservative than the code lists used for COVER. However, both data sources include codes which are acceptable with regards to payment for vaccination using the national business rules for general practice vaccination programmes, but also allow for wider codes related to the MMR vaccine.

**Supplementary Table S2. Comparison of yearly coverage estimates for second dose the MMR vaccine at age 5**

| **Year** | **MMR vaccine coverage in CPRD** | **National estimates for MMR in COVER** |
| --- | --- | --- |
| 2009-10 | - | 82.7 |
| 2010-11 | - | 84.2 |
| 2011-12 | 90.0 | 86.0 |
| 2012-13 | 90.8 | 87.7 |
| 2013-14 | 91.6 | 88.3 |
| 2014-15 | 91.2 | 88.6 |
| 2015-16 | 90.5 | 88.2 |
| 2016-17 | 89.9 | 87.6 |
| 2017-18 | 89.3 | 87.2 |
| 2018-19 | 89.2 | 86.4 |

Supplementary Figure S1. Vaccine coverage by data source and region – North East, North West and South East. Red lines represent data from COVER, green lines are data from COVER adjusted for 50% under-ascertainment and blue lines represent data from CPRD.

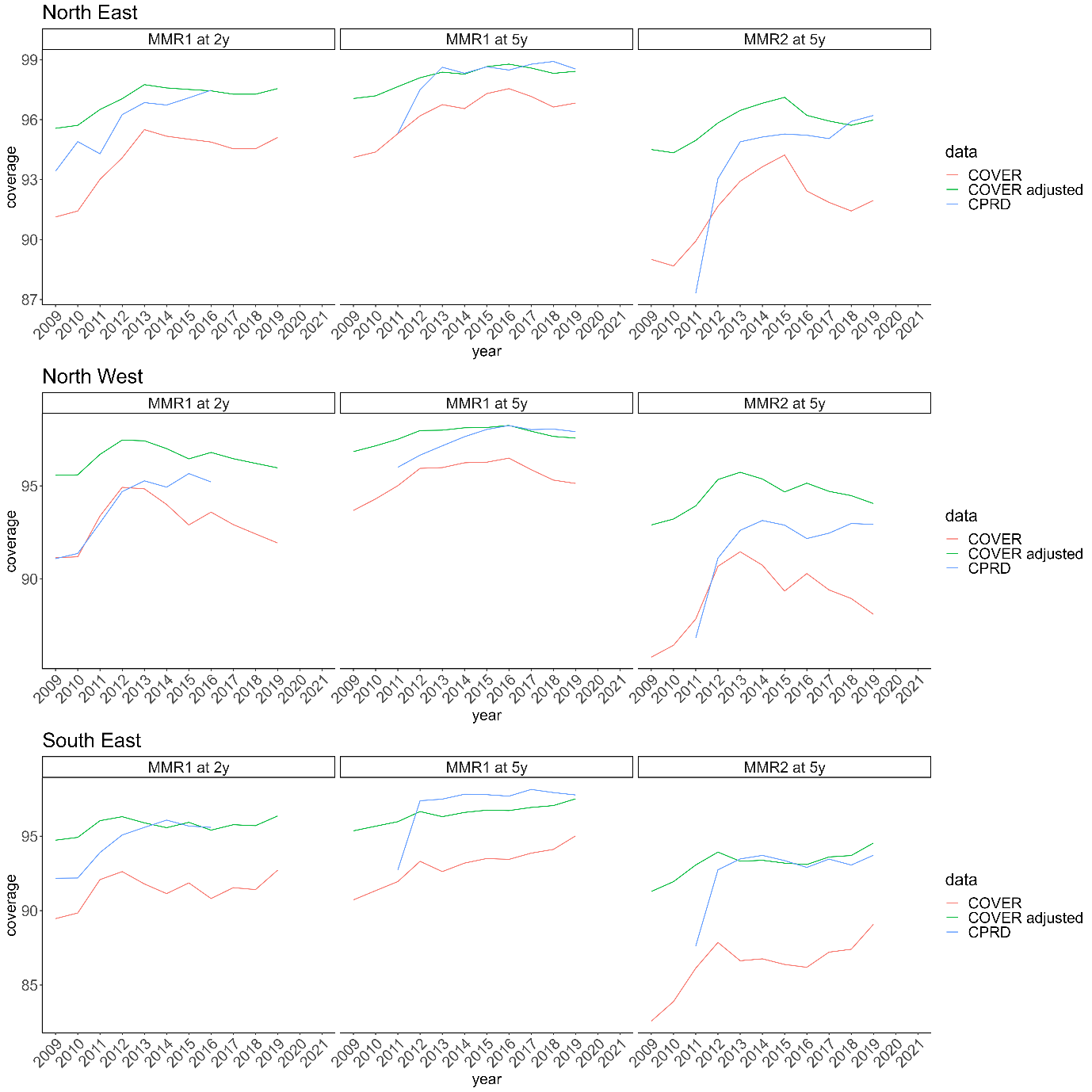

Supplementary Figure S2: Vaccine coverage by data source and region – South West, West Midlands and Yorkshire and the Humber. Red lines represent data from COVER, green lines are data from COVER adjusted for 50% under-ascertainment and blue lines represent data from CPRD.

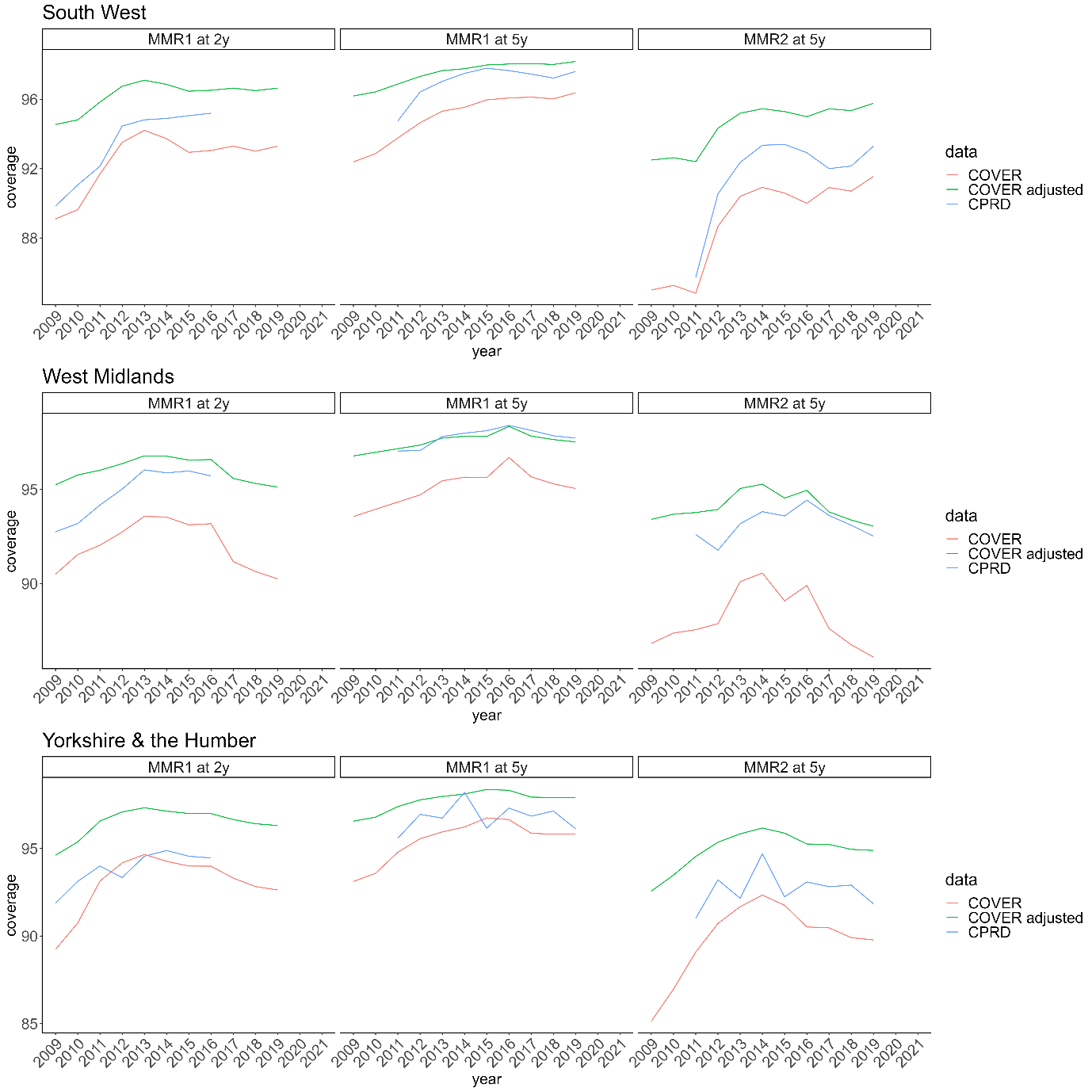

#### Data extrapolation in reference scenario using CPRD data

Data for age bands not included in CPRD (e.g., children born before 2006 for older age bands or after 2015 for children in the younger age bands), were supplemented with estimated values based on COVER data but adjusted for 50% under-ascertainment(9). This was done using the COVER estimates of the respective years and regions with an addition of 50% of the unvaccinated population ($original value + 0.5 * (100-original value)).$If information on specific age bands was missing due to the lower granularity of COVER, the supplemented value (i.e., five year coverage) was multiplied by the rate ratio of the five and four year coverage of the closest birth cohort with available data. The three year coverage was estimated accordingly.

#### Data extrapolation in sensitivity analysis using COVER data

The COVER data is not stratified by individual age band (only available at 2 and 5 years old). The missing values were extrapolated using the CPRD data. The missing age bands were derived by multiplying the 5 year coverage by the rate ratio of the five and four year coverage from the same birth cohort available in CPRD. Similarly, the supplemented four year coverage was also multiplied with the rate ratio of the four and three year coverage in order to estimate the three year coverage.

### Section S3: Model fit and stochastic simulations: reference scenario (using CPRD data)

#### Description of the model fit

All models have three stratifications: by region, age, and vaccine status. All model converged and parameters were clearly identified (Figure S3, S4, S5, and Table S3). The log-posterior distributions of all models showed that models with waning performed between than the model that did not include waning of vaccine-induced immunity.

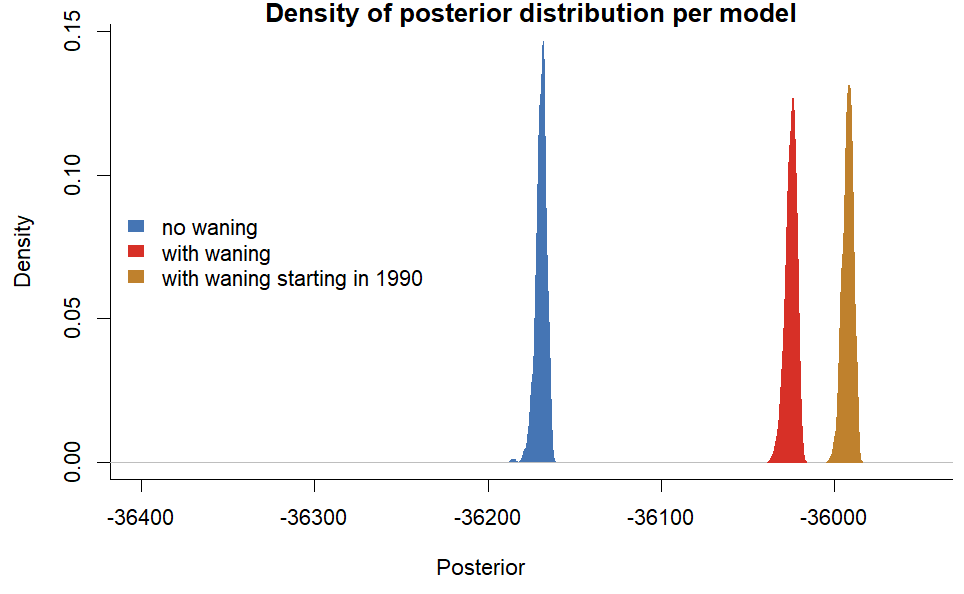

Supplementary Figure S3: Log-posterior density of each model in the reference scenario. Models that include waning performed better than the model that did not (higher posterior).

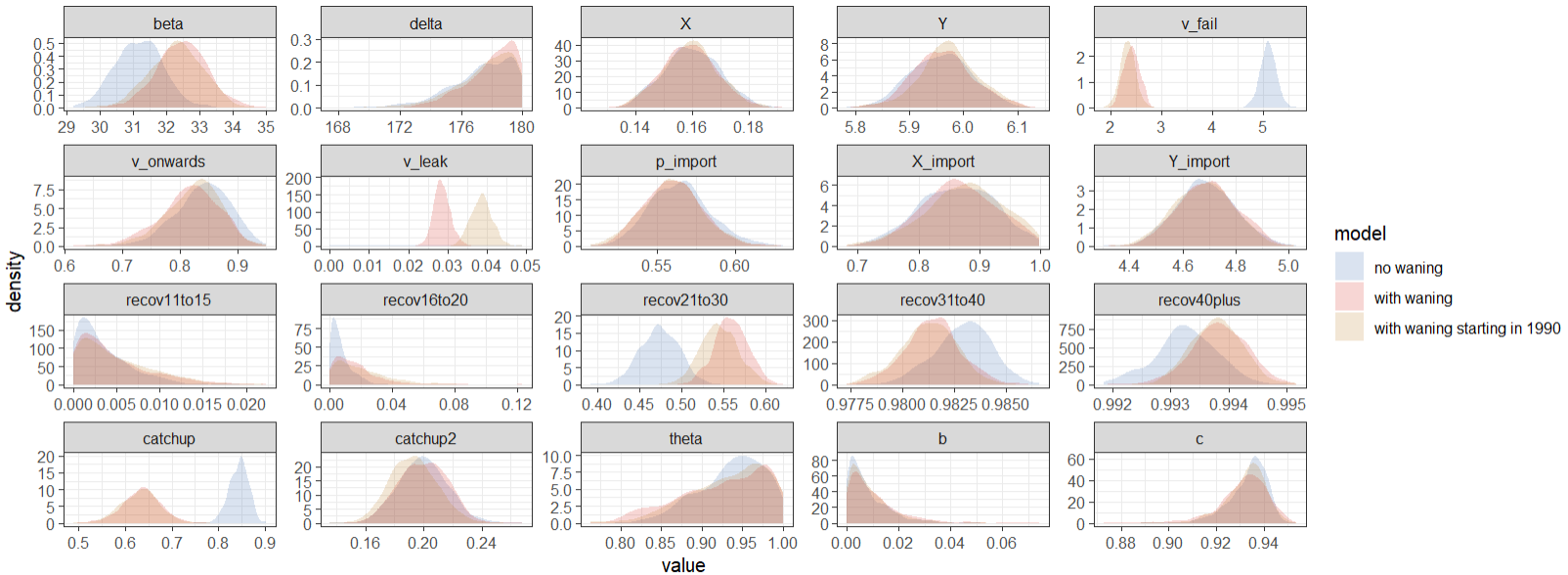

Supplementary Figure S4: Comparison of the posterior distribution of the estimated parameters for each model.

Supplementary Table S3: Parameter estimates in each model (Median and 95% credible intervals in brackets).

| Parameter | Prior | Without waning | With waning | With waning starting in 1990 |
| --- | --- | --- | --- | --- |
| Infection rate $\beta$ | U(0, 50) | 31 (30 - 33) | 33 (31 - 34) | 32 (31 - 34) |
| Duration of maternal immunity (days) $\delta$ | N(90, 5) | 178 (172 - 180) | 179 (174 - 180) | 178 (173-180) |
| Risk of onwards transmission from vaccinated cases compared to unvaccinated cases $v_{onwards}$ | Beta(10,10) | 0.84 (0.74 - 0.92) | 0.82 (0.71 - 0.9) | 0.83 (0.72 - 0.91) |
| Seasonality of infection rate: $X$ | U(0, 7) | 0.16 (0.14 - 0.18) | 0.16 (0.14 - 0.18) | 0.16 (0.14 - 0.18) |
| Seasonality of infection rate: $Y$ | U(0, 7) | 5.96 (5.85 - 6.07) | 5.96 (5.86 - 6.08) | 5.97 (5.86 - 6.08) |
| Waning of immunity per year $v_{leak}$ | U(0,1) | 0 | 2.9 (2.5 - 3.3) e-4 | 3.9 (3.4 – 4.4) e-4 |
| Proportion of primary vaccine failure $v_{fail}$ | U(0, 1) | 0.05 (0.05 - 0.05) | 0.02 (0.02 - 0.03) | 0.02 (0.02 - 0.03) |
| Proportion individuals vaccinated during the 1996 catch-up campaign $catchup$ | U(0, 1) | 0.85 (0.8 - 0.88) | 0.64 (0.56 - 0.71) | 0.63 (0.55 - 0.71) |
| Proportion individuals vaccinated during the 2008 catch-up campaign $catchup2$ | U(0, 1) | 0.2 (0.17 - 0.23) | 0.2 (0.17 - 0.23) | 0.19 (0.16 - 0.23) |
| Proportion of previously infected 10-15 yo $recov_{10-15}$ | U(0, 1) | 0 (0 - 0.01) | 0 (0 - 0.02) | 0 (0 - 0.02) |
| Proportion of previously infected 15-20 yo $recov_{15-20}$ | U(0, 1) | 0.01 (0 - 0.02) | 0.01 (0 - 0.06) | 0.02 (0 - 0.06) |
| Proportion of previously infected 20-30 yo $recov_{20-30}$ | U(0, 1) | 0.47 (0.43 - 0.52) | 0.56 (0.52 - 0.6) | 0.54 (0.5 - 0.59) |
| Proportion of previously infected 30-40 yo $recov_{30-40}$ | U(0, 1) | 0.98 (0.98 - 0.99) | 0.98 (0.98 - 0.98) | 0.98 (0.98 - 0.98) |
| Proportion of previously infected 40+ yo $recov_{40+}$ | U(0, 1) | 0.99 (0.99 - 0.99) | 0.99 (0.99 - 0.99) | 0.99 (0.99 - 0.99) |
| Spatial parameter $b$ | U(0, 5) | 0.01 (0 - 0.03) | 0.01 (0 - 0.04) | 0.01 (0 - 0.03) |
| Spatial parameter $c$ | U(0, 5) | 0.94 (0.92 - 0.94) | 0.93 (0.91 - 0.95) | 0.93 (0.91 - 0.95) |
| Spatial parameter $\theta$ | U(0, 1) | 0.94 (0.85 - 1) | 0.93 (0.81 - 1) | 0.94 (0.83 - 1) |
| Proportion of importations reported $p_{import}$ | U(0, 1) | 0.56 (0.53 - 0.61) | 0.56 (0.53 - 0.6) | 0.56 (0.52 - 0.6) |
| Seasonality of importations $X_{import}$ | U(0, 7) | 0.86 (0.74 - 0.97) | 0.87 (0.75 - 0.98) | 0.88 (0.74 - 0.98) |
| Seasonality of importations $Y_{import}$ | U(0, 7) | 4.69 (4.47 - 4.9) | 4.69 (4.48 - 4.9) | 4.68 (4.46 - 4.89) |

The comparison between parameter estimates in models with and without waning shows that most parameter estimates are similar in all models (Figure S5 and Supplementary Table S3). In all models, the proportion of individuals who had previously been infected with measles as of 2010 increased by age (Figure S5A). When using the COVER data (Supplementary Section S5), R0 was smaller (between 9.5 and 11) as more individuals start off as susceptible since the average vaccine coverage was lower in the COVER data. The duration of maternal immunity in all models was above the prior assumed distribution (Figure 5 Panel D: the estimated duration was 175 days, the mean of the prior is 90). Immunity in individuals below 1 only comes from maternal immunity. The POLYMOD study does not distinguish between 3-5 year-olds and infants, likely leading to an overestimation of contacts in children below 1.

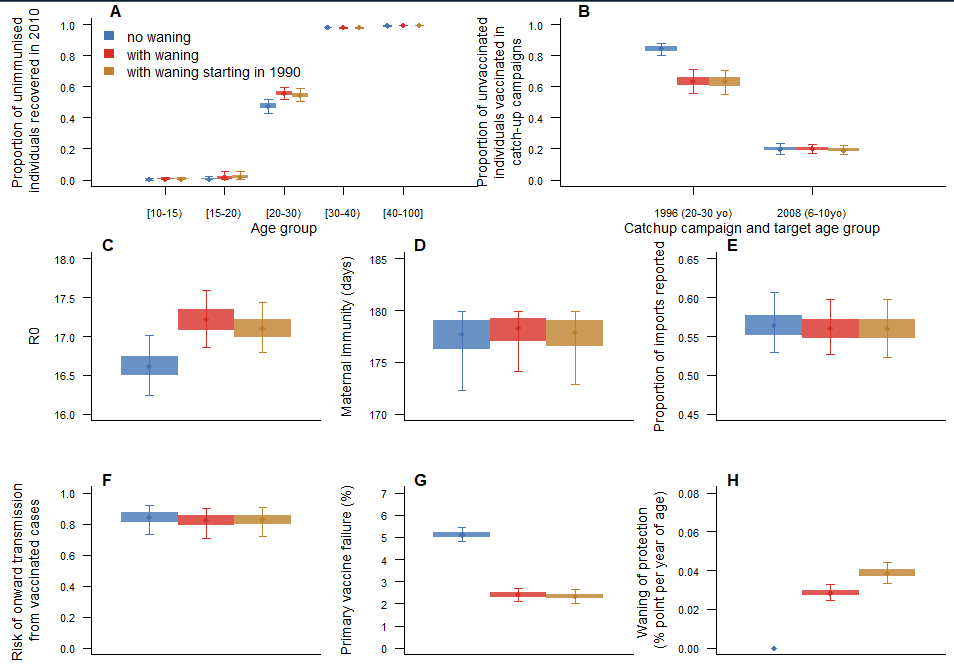

Supplementary Figure S5: Parameter estimates in models with and without waning of vaccine-induced immunity (using CPRD vaccine data). A. Proportion of unvaccinated individuals who start off as recovered per age group. B. Proportion of susceptible individuals who were not vaccinated during routine vaccination campaigns, but got vaccinated during catchup up campaigns before 2010 (the 1996 campaign targeted individuals aged 20 to 30 in 2010, the 2008 campaign targeted individuals aged 6 to 10 in 2010). C. R0 in each model. D. Duration of maternal immunity in days. E. Proportion of importations reported. F. Risk of onward infection in vaccinated cases compared to unvaccinated cases. G. Percentage of primary vaccine failure. H. Rate of waning of vaccine-induced immunity (in percentage point per year of age).

The number of cases stratified by region and age follow a similar distribution, independent of the presence of waning immunity (Figure S6). The improvements in posterior distribution therefore mostly stem from changes in age-distribution and timing of vaccinated cases (Figure 3 and 4).

The distribution of the number of cases by age groups is in agreement with the data. When using CPRD data, the model over-estimates the number of infants reported (below one year old), this is due to the number of contacts among infants, and the fact that they have no prior immunity. This is especially visible in the proportion of cases per age group (Figure S6 Panel D).

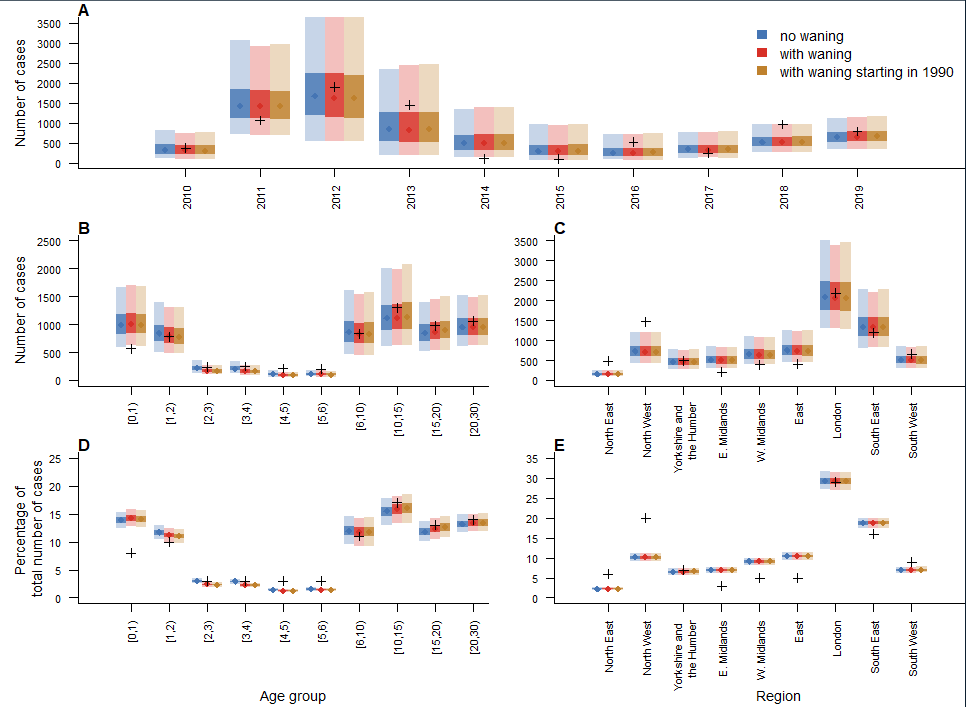

Supplementary Figure S6: A. Number of cases per year in each model and in the data across all regions and age groups (black crosses). B. Number of cases by age groups in the models and the data across all regions and years. C. Number of cases by regions in the models and the data across all regions and years. D. Proportion of cases by age groups in the models and the data. E. Proportion of cases by regions in the models and the data.

The seasonality of the transmission parameters and number of importations is presented in Figure S7. Transmission risk is highest between December and February, and lowest in June to August. Importation rates are highest between March and May, which are the months where most cases were reported.

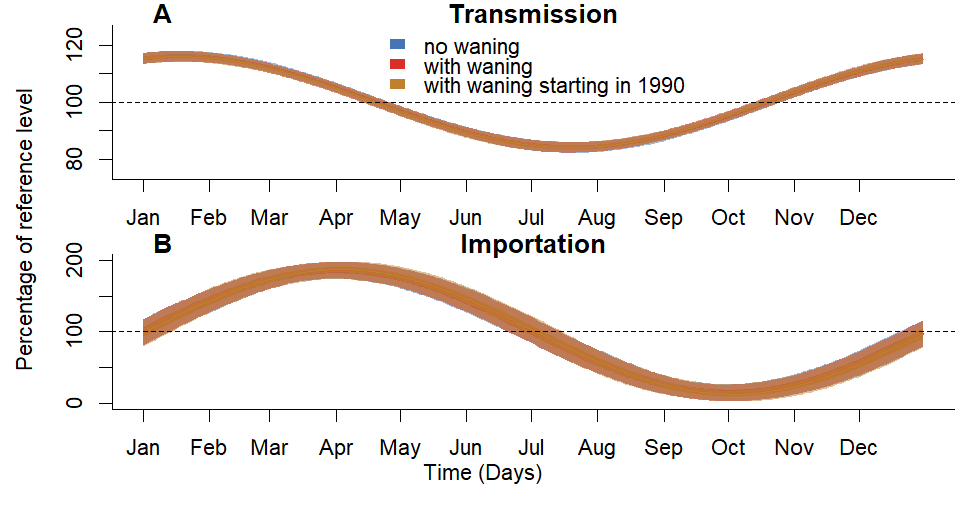

Supplementary Figure S7: Within-year seasonality of transmission (A) and importation (B) parameters. All models fits lead to similar seasonality.

#### Number of cases caused by waning (reference scenario, using CPRD data)

In the reference scenario, the proportion of cases averted by removing waning was high, especially in 2018 and 2019 (Figure S8). This reduction is due to both direct effects (vaccinated cases were only due to primary vaccine failures, which were rarer than in models without waning of immunity) and indirect effects (unvaccinated cases infected by vaccinated cases with waning were not infected when waning was removed).

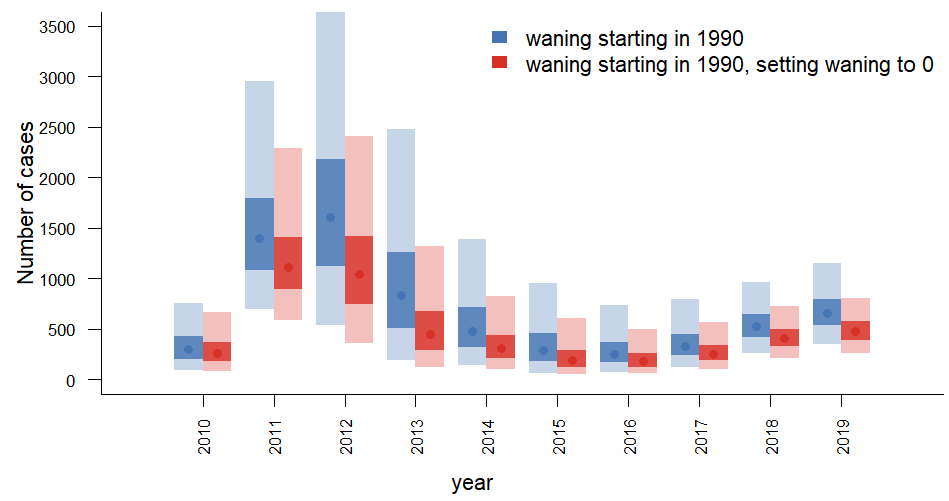

Supplementary Figure S8: Number of yearly cases in simulations generated using the model with waning starting in 1990, and after setting the waning parameter to 0.

### Section S4: Model fit and stochastic simulations with baseline risk of secondary vaccine failure

When a constant risk of secondary vaccine failure was added to the reference scenarios, the model without waning of vaccine-induced immunity captures the number of single and double vaccinated in the whole as well as models that included waning (Figure S9, Panel A). However, simulations from the model without waning did not capture the age distribution of double-vaccinated cases, overestimating the number of double-vaccinated cases under 15 (Median 150 [95% simulation interval (SI): 85-265] cases, 66 cases in the data), and underestimating the number of double-vaccinated teenagers and adults (Median 135 [95%SI: 84-219] cases, 202 cases in the data) (Figure S9, panels C-D-F-G).

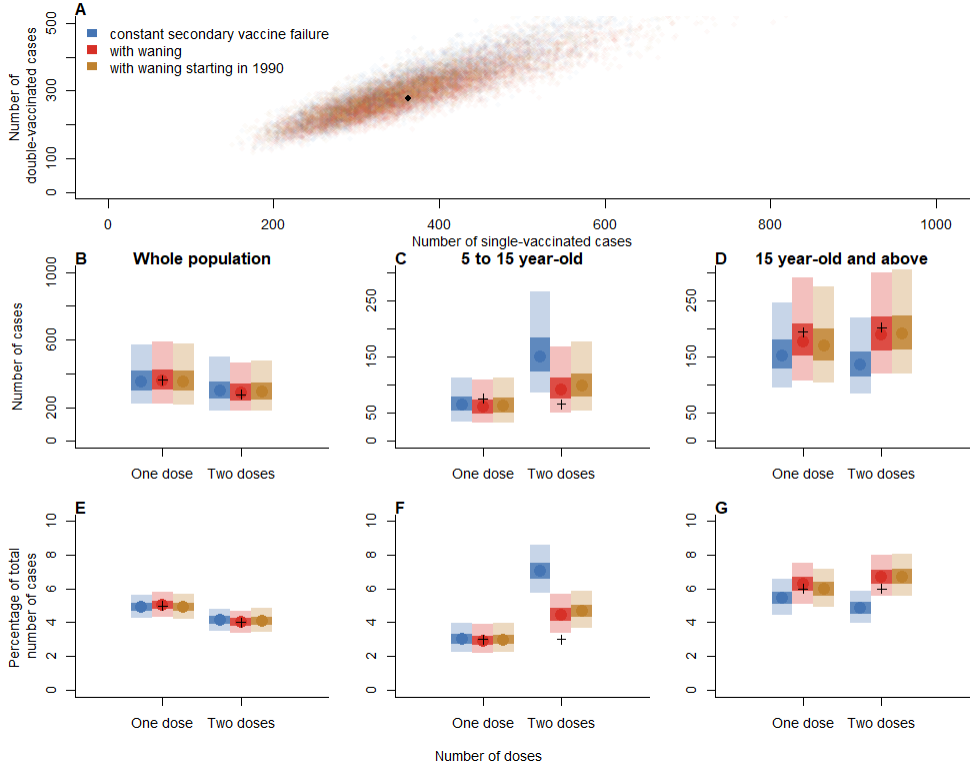

Supplementary Figure S9: A. Comparison of the number of single and double vaccinated cases in models with and without waning. B. Overall number of single and double vaccinated cases in each model (data points are represented by black crosses). C. Number of vaccinated cases between 5 and 15 years old. D. Number of vaccinated cases above 15 years old. E. Overall proportion of single and double vaccinated cases. F. Proportion of vaccinated cases between 5 and 15 years old. G. Number of vaccinated cases above 15 years old.

As in the reference scenario, only models that included waning of immunity captured the increase in proportion of double-vaccinated cases observed in the data between 2010 and 2019 (Figure S10). In the model with constant risk of secondary vaccine failure, the median proportion of double-vaccinated cases was constant around 4% each year.

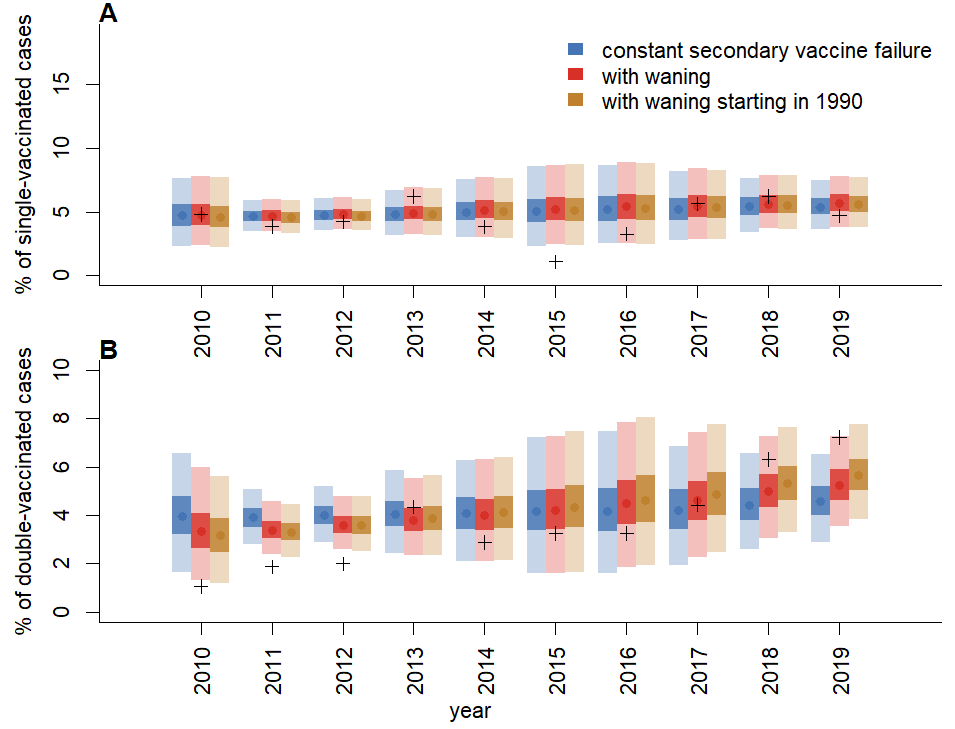

Supplementary Figure S10: A. Proportion of single (and B. double) vaccinated cases each year across all regions and age groups.

When a risk of secondary vaccine failure constant with age was added to the model, the parameter estimates were similar to the reference scenario (Figure S11). The parameters of the model without waning were now more similar to models that incorporated waning (i.e. similar primary vaccine failure and R0). However, the baseline risk of secondary vaccine failure in models that include waning was almost 0, all secondary vaccine failure was explained by the waning of immunity.

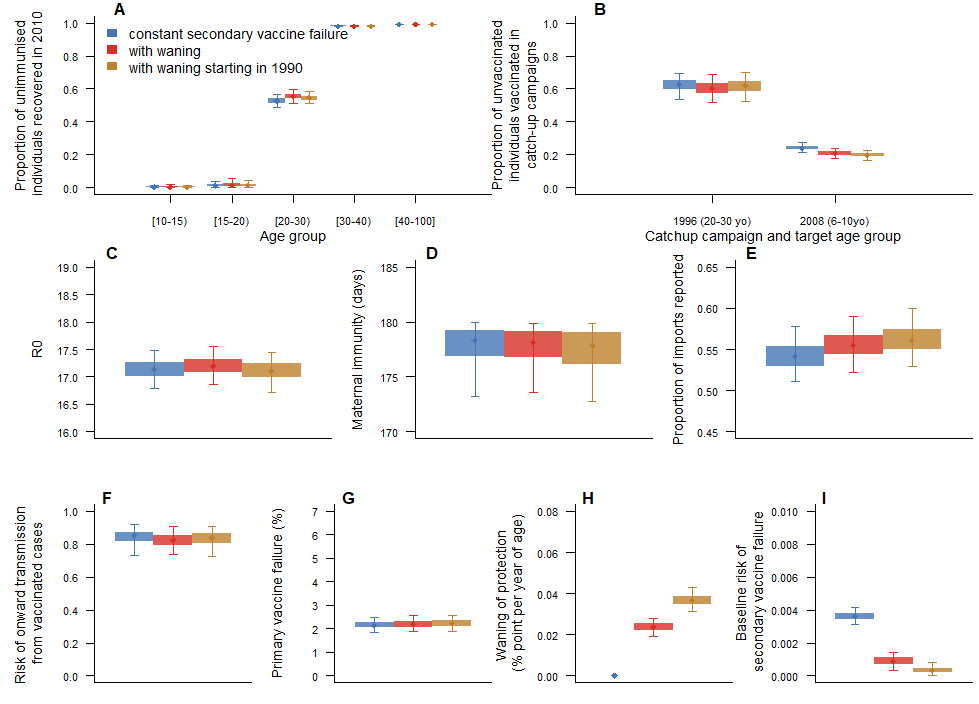

Supplementary Figure S11: Parameter estimates in models with and without waning of vaccine-induced immunity (using CPRD vaccine data, with baseline risk of secondary vaccine failure). A. Proportion of unvaccinated individuals who start off as recovered per age group. B. Proportion of susceptible individuals who were not vaccinated during routine vaccination campaigns, but got vaccinated during catchup up campaigns before 2010 (the 1996 campaign targeted individuals aged 20 to 30 in 2010, the 2008 campaign targeted individuals aged 6 to 10 in 2010). C. R0 in each model. D. Duration of maternal immunity in days. E. Proportion of importations reported. F. Protection against onward infection brought by the vaccine. G. Percentage of primary vaccine failure. H. Rate of waning of vaccine-induced immunity (in percentage point per year of age). I) Baseline risk of secondary vaccine failure.

The model fits were similar to the reference scenario: the age distribution in the simulation was similar to the data, but bigger discrepancies were observed in the spatial distribution. The distributions were similar for all three models (Figure S12).

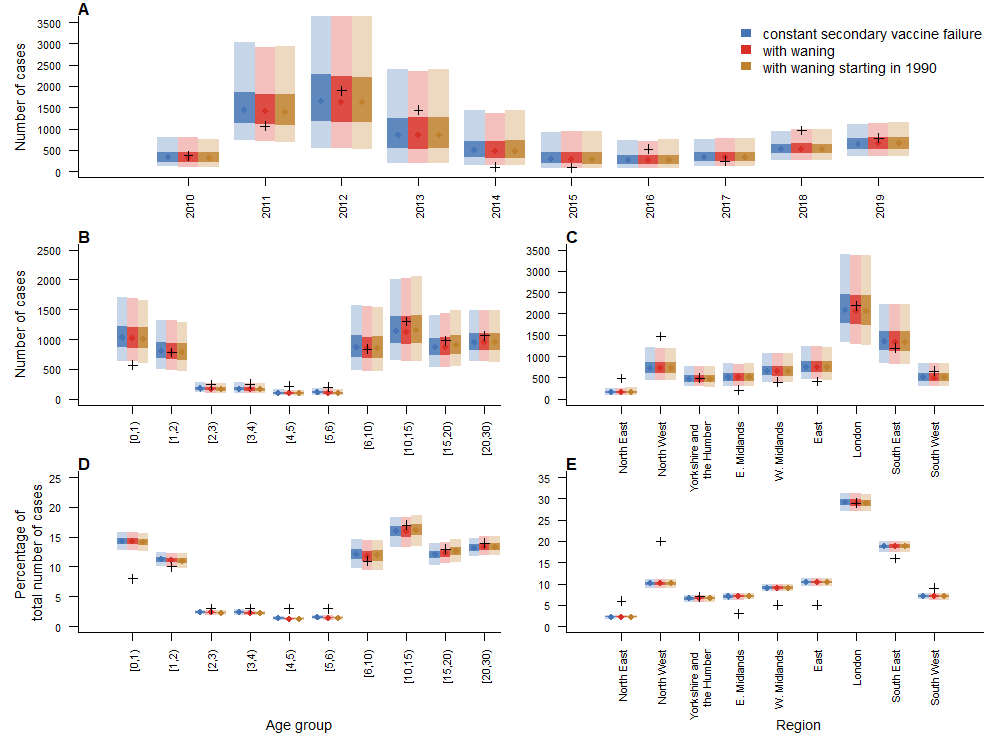

Supplementary Figure S12: A. Number of cases per year in each model and in the data across all regions and age groups (black crosses). B. Number of cases by age groups in the models and the data across all regions and years. C. Number of cases by regions in the models and the data across all regions and years. D. Proportion of cases by age groups in the models and the data. E. Proportion of cases by regions in the models and the data.

### Section S5: Model fit and stochastic simulations using COVER data

Similar to the reference scenario, fitting the model using COVER data showed that the number of single and double vaccinated cases could only be captured by models that included waning of immunity (Figure S13 Panel A). If only primary vaccine failure was considered, the simulations did not capture the age distribution and number of vaccinated cases (Figure S13).

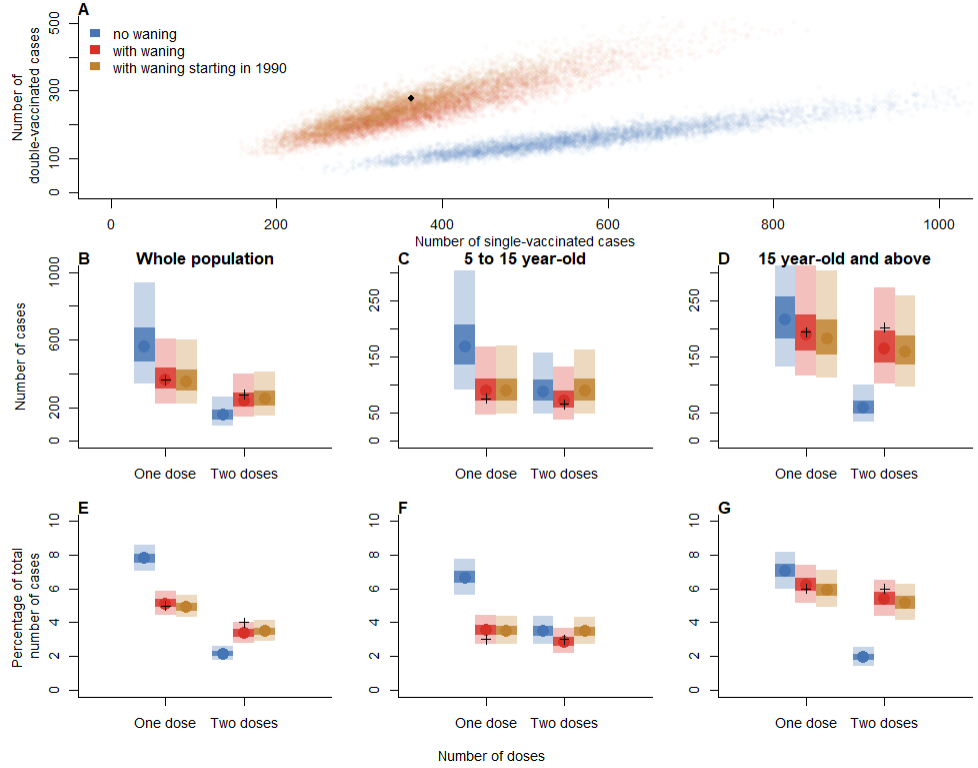

Supplementary Figure S13: A. Comparison of the number of single and double vaccinated cases in models with and without waning. B. Overall number of single and double vaccinated cases in each model (data points are represented by black crosses). C. Number of vaccinated cases between 5 and 15 years old. D. Number of vaccinated cases above 15 years old. E. Overall proportion of single and double vaccinated cases. F. Proportion of vaccinated cases between 5 and 15 years old. G. Number of vaccinated cases above 15 years old.

Only the models with waning of immunity showed an increase in the proportion of double vaccinated cases through time (Figure S14), although this increase was slower than in the reference scenario, and in the data.

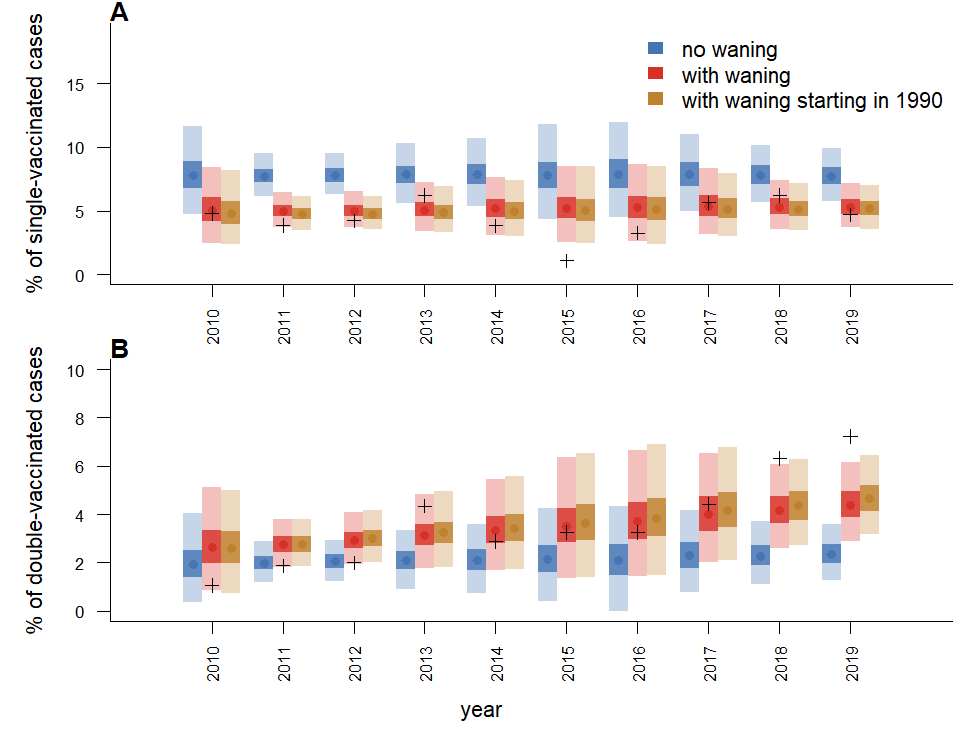

Supplementary Figure S14: A. Proportion of single (and B. double) vaccinated cases each year across all regions and age groups.

The model fits show that all models capture the number of cases younger than 1 better than the CPRD scenario, although the simulations under estimate the number of cases aged 1 to 2. As in the CPRD scenario, there are discrepancies in the spatial distribution of the cases between the simulations and the data (Figure S15).

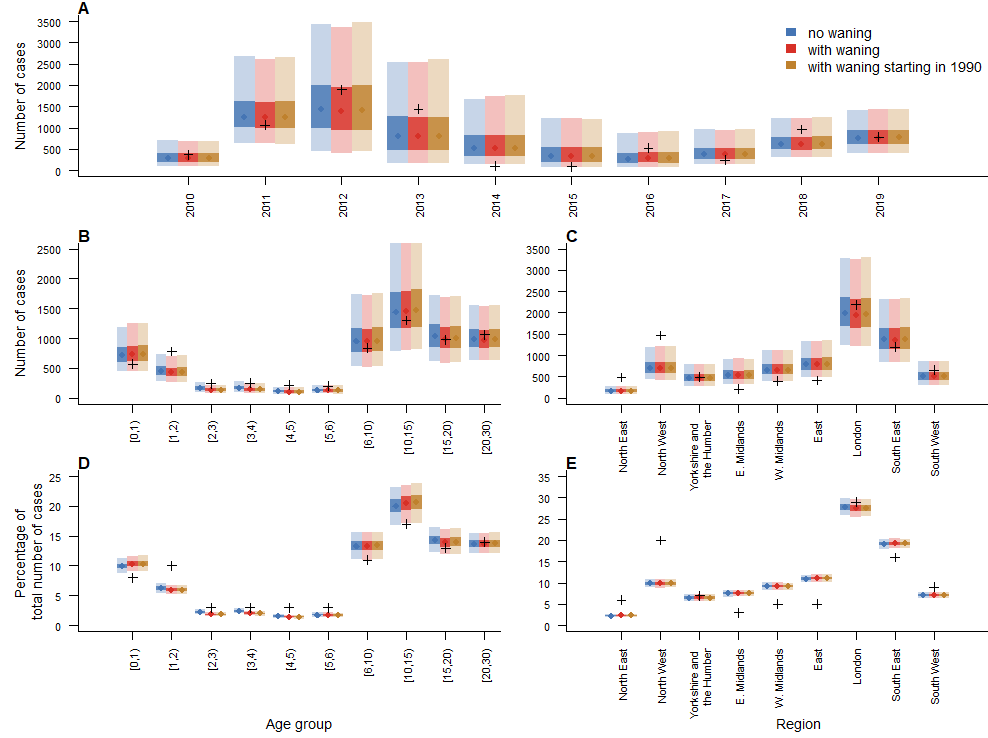

Supplementary Figure S15: A. Number of cases per year in each model and in the data across all regions and age groups (black crosses). B. Number of cases by age groups in the models and the data across all regions and years. C. Number of cases by regions in the models and the data across all regions and years. D. Proportion of cases by age groups in the models and the data. E. Proportion of cases by regions in the models and the data.

In the models ran using the COVER data, the duration of maternal immunity followed the prior distribution, as the proportion of vaccinated individuals is lower than in the CPRD data (Figure S16). The basic reproduction number was also lower than in the reference scenario. Both differences are due to the lower proportion of vaccinated individuals in the COVER data compared to the CPRD scenario.

The risk of onwards transmission from vaccinated cases was lower in the COVER scenario than in the reference scenario (Median: 21%, 95% credible interval (CI): 11-35%)). This means that vaccinated cases were less likely to cause onwards transmission than unvaccinated cases. This may be due to the lower level of immunisation in the community, where simulated outbreaks would get too big if vaccinated cases were transmitting as much as unvaccinated cases. As a consequence, removing waning from the scenario where waning started in 1990 had less impact than in the reference scenario (Figure S17), as most averted cases were due to direct effects (vaccinated cases not being infected if secondary vaccine failure is removed from the model).

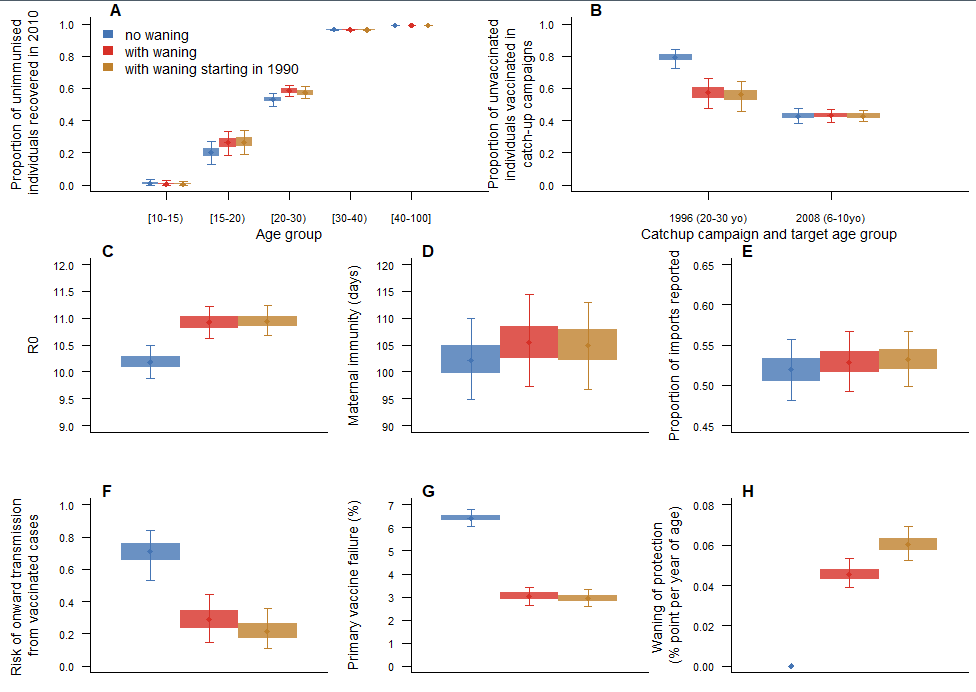

Supplementary Figure S16: Parameter estimates in models with and without waning of vaccine-induced immunity (using COVER vaccine data). A. Proportion of unvaccinated individuals who start off as recovered per age group. B. Proportion of susceptible individuals who were not vaccinated during routine vaccination campaigns, but got vaccinated during catchup up campaigns before 2010 (the 1996 campaign targeted individuals aged 20 to 30 in 2010, the 2008 campaign targeted individuals aged 6 to 10 in 2010). C. R0 in each model. D. Duration of maternal immunity in days. E. Proportion of importations reported. F. Risk of onward infection in vaccinated cases compared to unvaccinated cases. G. Percentage of primary vaccine failure. H. Rate of waning of vaccine-induced immunity (in percentage point per year of age).

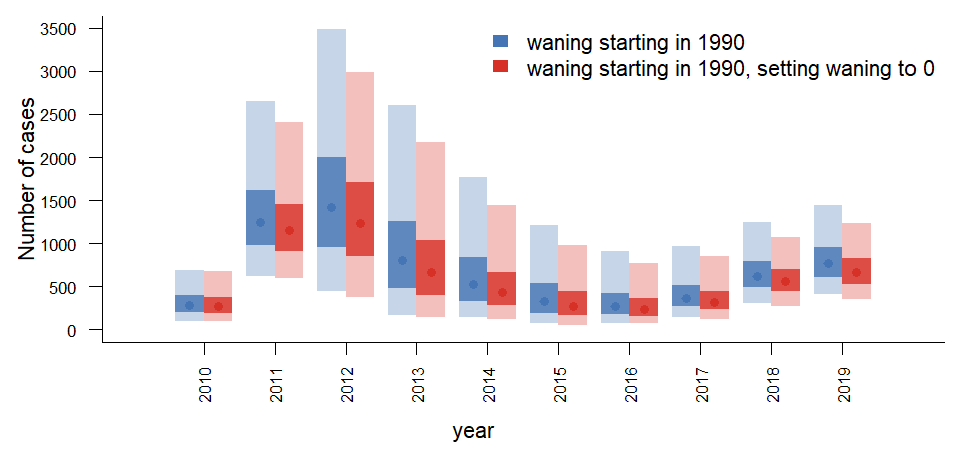

Supplementary Figure S17: Number of yearly cases in simulations generated using the model with waning starting in 1990,, and after setting the waning parameter to 0.

### Section S6: Model fit and stochastic simulations with fixed distance kernel

Is this scenario, the parameters of the spatial were set ($theta=1$, $b=1$, $c=1$ in Table S2). We implemented this scenario to explore the impact of changing the movements between regions on the estimates of the model, in particular whether that changed the distribution of vaccinated cases in the simulation.

The results were very similar to that of the reference scenario: only models that incorporated waning of immunity could capture the age and time distribution of vaccinated cases. Models that did not consider any secondary vaccine failure over estimated the number of single vaccinated cases, and underestimated the number of double-vaccinated teenagers and young adults who got infected (Figure S18). Only models that incorporated waning of immunity showed an increase in the proportion of double vaccinated cases through time (Figure S19).

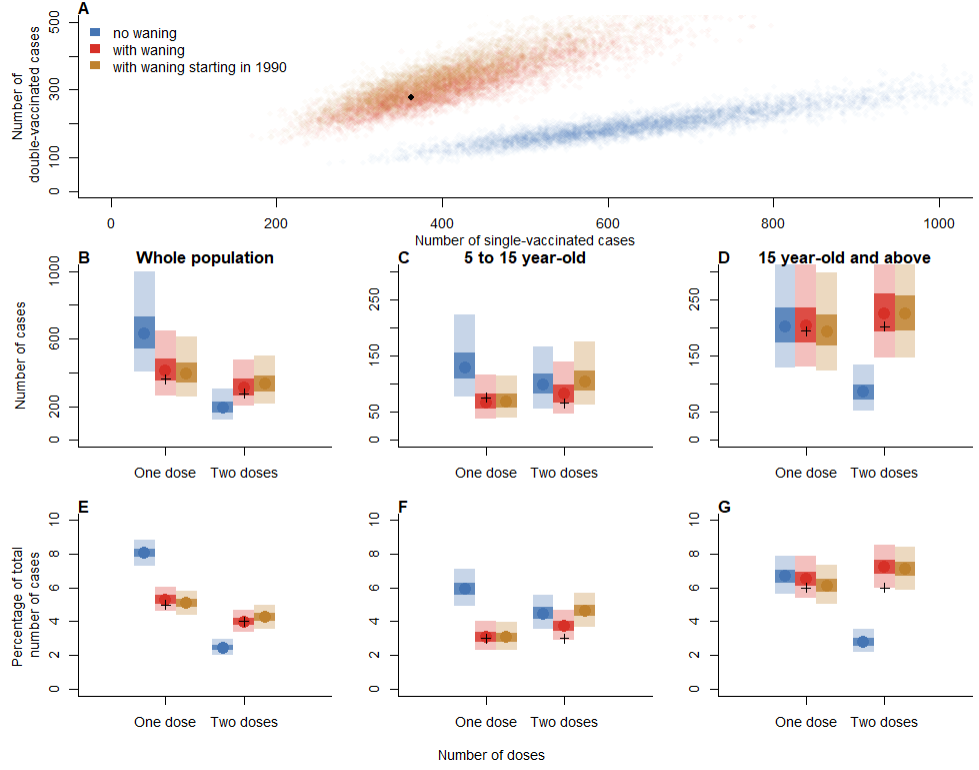

Supplementary Figure S18: A. Comparison of the number of single and double vaccinated cases in models with and without waning. B. Overall number of single and double vaccinated cases in each model (data points are represented by black crosses). C. Number of vaccinated cases between 5 and 15 years old. D. Number of vaccinated cases above 15 years old. E. Overall proportion of single and double vaccinated cases. F. Proportion of vaccinated cases between 5 and 15 years old. G. Number of vaccinated cases above 15 years old.

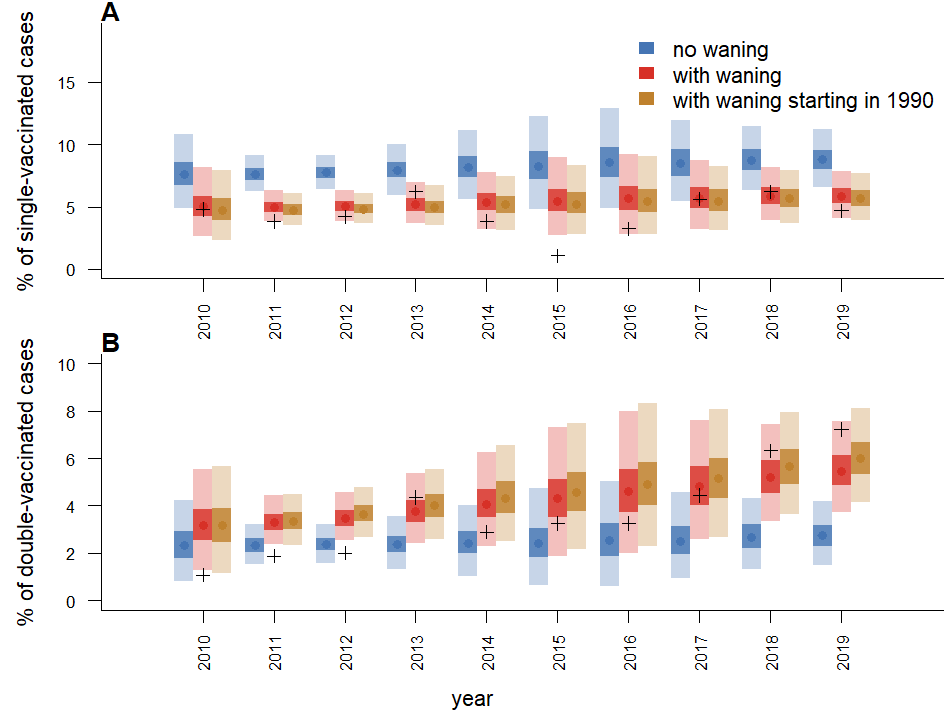

Supplementary Figure S19: A. Proportion of single (and B. double) vaccinated cases each year across all regions and age groups.

The parameter estimates and age-distribution of the cases was the same as the reference scenario (Figures S20 and S21), while the spatial distribution was slightly worse than the reference scenario as the number of cases in South East was over estimated in this scenario.

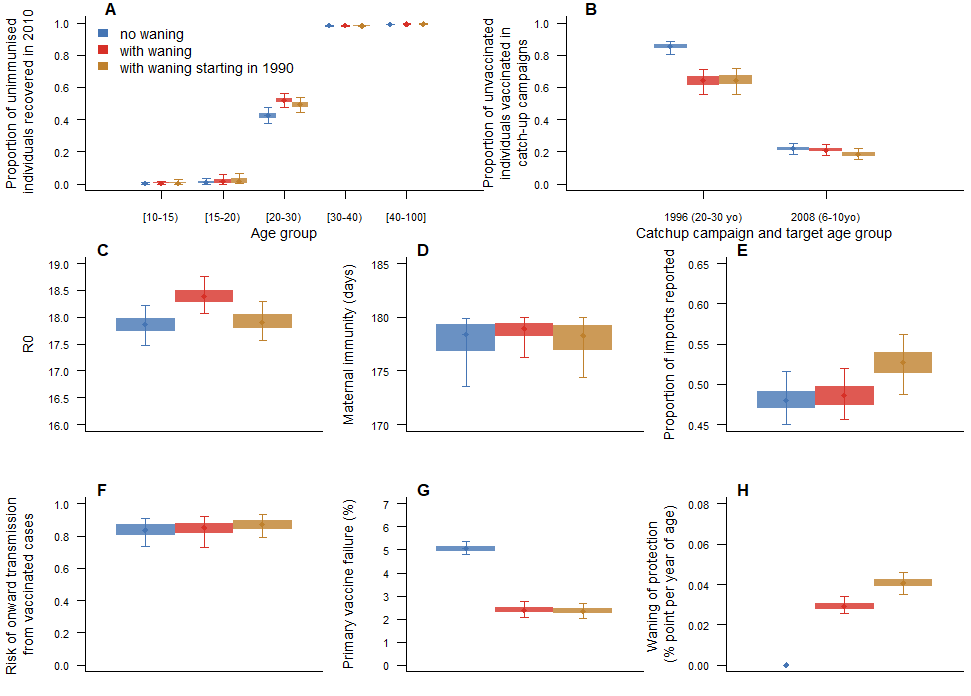

Supplementary Figure S20: Parameter estimates in models with and without waning of vaccine-induced immunity (using CPRD vaccine data, with a fixed spatial kernel). A. Proportion of unvaccinated individuals who start off as recovered per age group. B. Proportion of susceptible individuals who were not vaccinated during routine vaccination campaigns, but got vaccinated during catchup up campaigns before 2010 (the 1996 campaign targeted individuals aged 20 to 30 in 2010, the 2008 campaign targeted individuals aged 6 to 10 in 2010). C. R0 in each model. D. Duration of maternal immunity in days. E. Proportion of importations reported. F. Risk of onward infection in vaccinated cases compared to unvaccinated cases. G. Percentage of primary vaccine failure. H. Rate of waning of vaccine-induced immunity (in percentage point per year of age).

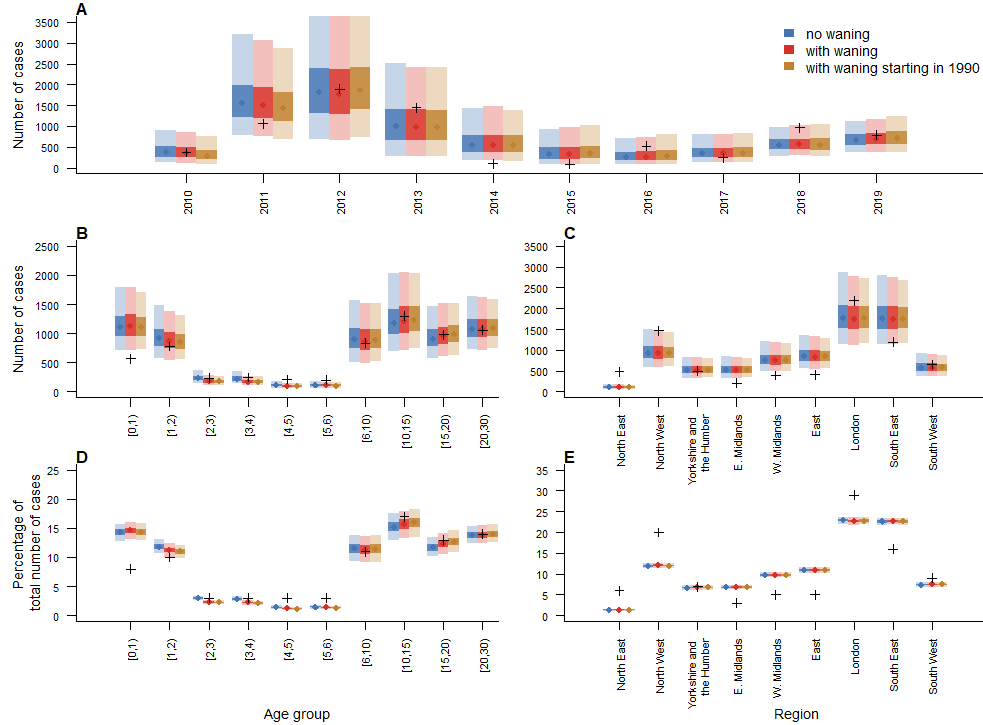

Supplementary Figure S21: A. Number of cases per year in each model and in the data across all regions and age groups (black crosses). B. Number of cases by age groups in the models and the data across all regions and years. C. Number of cases by regions in the models and the data across all regions and years. D. Proportion of cases by age groups in the models and the data. E. Proportion of cases by regions in the models and the data.
